# Efficacy of Naloxone in reducing hypoxemia and duration of immobility following focal to bilateral tonic-clonic seizures

**DOI:** 10.1101/2024.12.16.24319102

**Authors:** Sylvain Rheims, Fatima Chorfa, Véronique Michel, Edouard Hirsch, Louis Maillard, Luc Valton, Fabrice Bartolomei, Philippe Derambure, Vincent Navarro, Julien Biberon, Arielle Crespel, Anca Nica, Martine Lemesle Martin, Laure Mazzola, Jerome Petit, Vincent Rossero, Sébastien Boulogne, Mathilde Leclercq, Laurent Bezin, Catherine Mercier, Pascal Roy, Philippe Ryvlin, the ENALEPSY study group

**Affiliations:** Department of Functional Neurology and Epileptology, Hospices Civils de Lyon and University of Lyon, France; Lyon’s Neuroscience Research Center, INSERM U1028 / CNRS UMR 5292, Lyon, France; Department of Biostatistics, Hospices Civils de Lyon and University of Lyon, France; Department of Neurology, HoCpital Pellegrin, Bordeaux, France; Department of Neurology, University Hospital of Strasbourg, Strasbourg, France; Neurology Department, University Hospital of Nancy, Nancy, France; Department of Neurology, University Hospital of Toulouse, & Brain and Cognition Research Centre (CerCo), CNRS, UMR5549, Toulouse, France; APHM, Timone Hospital, Epileptology and Cerebral Rhythmology, Marseille, France; Department of Clinical Neurophysiology, Lille University Medical Center, EA 1046, University of Lille2; AP-HP, Epilepsy Unit, Pitié-Salpêtrière Hospital, Sorbonne Université, and Paris Brain Institute (ICM; INSERM, CNRS) ERN EpiCare, Paris, France; Department of Clinical Neurophysiology, University Hospital of Tours, Tours, France; Epilepsy Unit, Montpellier, France; Department of Neurology, University Hospital of Rennes, Rennes, France; Department of Neurology, University Hospital of Dijon, Dijon, France; Department of Neurology, University Hospital of Saint-Etienne, Saint-Etienne, France; La Teppe Epilepsy Center, Tain l’Hermitage, France; Department of Clinical Neurosciences, Centre Hospitalo-Universitaire Vaudois, Lausanne, Switzerland

**Author notes:** Correspondence to: Pr Sylvain Rheims, Department of Functional Neurology and Epileptology 59 boulevard Pinel, 69677 Bron cedex, France. The statistical analyses were conducted by Fatima Chorfa, Catherine Mercier and Pr Pascal Roy, Department of Biostatistics. **Trial registration**: ClinicalTrials.gov NCT02332447; EudraCT 2014-003003-30. **Funding:** The ENALEPSY study was funded by the French Ministry of Health (PHRC National 2013). The REPO_2_MSE study was funded by the French Ministry of Health (PHRC National 2009). Vincent Rossero is funded by ERANET-NEURON – Agence Nationale de la Recherche, Grant/ Award Number: ANR-21-NEU2-0006 Sylvain Rheims is funded by HORIZON-ERC-Award Number: ERC-2023-COG-101125118.

**Keywords:** epilepsy, SUDEP, post-ictal EEG hypoxemia, post-ictal immobility, generalized convulsive seizures, naloxone

## Abstract

**Purpose:** Evaluating the efficacy of an opioid antagonist, naloxone (NLX), to reduce the severity of post-ictal hypoxemia and immobility after focal to bilateral tonic-clonic seizures (FBTCS).

**Methods:** ENALEPSY is a double-blind placebo (PCB)-controlled trial conducted in patients with focal epilepsy undergoing long-term video-EEG monitoring (LTM). Patient with a FBTCS during LTM were randomized 1:1 to receive intravenous NLX or PCB within the 2 minutes following the end of FBTCS. After database lock, a discrepancy between the allocated arm and the received treatment was detected, resulting in a 4:1 NLX:PCB ratio. To further explore the efficacy of NLX, we used historical control (HC) data collected in patients included in the REPO_2_MSE study whose characteristics matched those of patients randomized in ENALEPSY. Efficacy of NLX was then assessed versus PCB and versus HC. The primary endpoint was the delay between the end of the seizure and recovery of SpO_2_ ≥ 90%. Secondary efficacy outcomes included desaturation nadir and duration of the postictal immobility.

**Results:** 33 patients contributed to the NLX group, 7 to the PCB group and 43 to the HC group. The proportion of FBTCS type 1 or 3 was 84% in NLX, 100% in PCB and 84% in HC. NLX did not improve the delay of recovery of SpO_2_ ≥ 90% or the desaturation nadir. In contrast, duration of the postictal immobility differed across groups. The time to mobility recovery within the first 5 minutes post-ictal was very similar in PCB (200.3±215.8 seconds) and HC (194.4±192.0 second) groups, and significantly shorter in the NLX group (128.9±151.1 seconds) when compared to HC (Hazard Ratio, 1.84; 95% CI, 1.11 to 3.05; p=0.021).

**Conclusion:** NLX did not prevent postictal respiratory dysfunction but might reduce the duration of postictal immobility. Confirmation of this effect and its impact on SUDEP risk will require additional studies

**KEY BULLET POINTS:**

– ENALEPSY trial evaluated the efficacy of intravenous naloxone to reduce the severity of post-ictal complications after focal to bilateral tonic-clonic seizures.
– Naloxone did not improve the delay of recovery of SpO_2_ ≥ 90% or the desaturation nadir
– The time to mobility recovery was significantly shorter in the NLX group when compared to historical control
– Impact of these results on SUDEP prevention will require additional studies

## INTRODUCTION

Currently, there is no effective treatment to prevent Sudden unexpected death in epilepsy (SUDEP)^(1)^, aside from optimizing antiseizure medications (ASM)^(1;2)^, epilepsy surgery or vagal nerve stimulation^(3)^. The exact mechanisms leading to SUDEP remain unknown ^(4)^, but central respiratory dysfunction is believed to play an important role ^(2; 4)^. Indeed, in most observed SUDEP cases, an apnea will first occur within 1-3 minutes after the termination of a generalized tonic-clonic seizures ^(5)^, followed by a terminal respiratory then cardiac arrest ^(5)^.

Several factors might contribute to post-ictal respiratory dysfunction, such as a brainstem spreading depolarization, but also possibly a release of endogenous opioid peptides^(6)^. Indeed, both animal and human studies have shown that such peptides are released during seizures, possibly to help aborting seizures^(7–9)^. Yet, given the rich concentration of opioid receptors in brainstem respiratory centers^(10)^, an excessive release of endogenous opioid peptides could also promote post-ictal respiratory dysfunction as observed in opioid overdose ^(11)^. If this was the case, opioid antagonists could offer a potential strategy to prevent SUDEP.

In order to test this hypothesis, we performed a proof-of-concept study that tested the acute effect of intravenous naloxone on postictal respiratory dysfunction and hypoxemia in patients with refractory focal epilepsy, when administered immediately after a focal to bilateral tonic-clonic seizures (FBTCS) during video-EEG monitoring.

## 2. METHODS

### 2.1 Study design

The ENALEPSY study (NCT02332447) is a multicenter, double blind, randomized placebo-controlled trial conducted in patients with focal epilepsy undergoing long-term video-EEG monitoring (LTM) in 15 epilepsy monitoring units in France (see Appendix 1 for the list of investigators and Appendix 2 for CONSORT checklist)^(12)^. Because patients who will develop a FBTCS during the LTM cannot be individualized a priori, we used a two-step design: all eligible patients were included in the study at the beginning of the LTM, equipped with a peripheral venous catheter and continuously monitored. Then, only those with a FBTCS during the LTM were randomized 1:1 to receive intravenous naloxone (NLX) or placebo (PCB) within the 2 minutes following the end of a FBTCS.

However, after completion of the planned recruitment, database lock and unblinding, a discrepancy between the allocated arm defined by randomization and the treatment actually received was detected in 20 of the 49 randomized patients. Specifically, 17 patients randomized to the PCB group actually received NLX whereas 3 randomized to the NLX group received PCB. Overall, while the randomization should have resulted in NLX administration in 24 patients and PCB in 25, 38 patients were eventually treated with NLX and 11 with PCB. The internal audit conducted by the coordination team (SR and ML), the biostatistics team (CM and PRoy) and the representants of the study sponsor (Hospices Civils de Lyon) revealed that this major protocol violation had resulted from a mismanagement of the treatment batches preparation by the pharmaceutical department of Hospices Civils de Lyon which had introduced a shift in the randomization. Given that the randomization was stratified by center and balanced by block (see below), this offset ultimately led to decorrelation with the randomization list and 4:1 NLX:PCB ratio

In this context, and prior to any statistical analysis of the data, the study steering committee (SR, PR, CM, PRoy and PI of each participating center) concluded that the number of subjects who received placebo was too low to offer an appropriate statistical power, and that the efficacy of NLX shall be further explored trough a comparison with an historical control (HC) group which did not received treatment, using the data collected in patients included in the REPO_2_MSE study and previously reported^(13)^. The rationale for this approach was the similarities of patients’ characteristics between REPO_2_MSE and ENALEPSY studies, which inclusion criteria and recruitment modalities were mostly similar. The original statistical plan was revised to integrate this combined analysis but without any modification of the primary or secondary endpoints which were analyzed in the HC group using the same approach as that predefined for the ENALEPSY trial.

### 2.2 Standard Protocol Approvals, Registrations, and Patient Consents”

Both ENALEPSY and REPOMSE were approved by ethics committee (CPP Sud Est II n°2014-853 and CPP Sud Est II n°2010-006-AM6) and competent authority (ANSM n° 141241A-31 and ANSM B100108-40) and all patients gave written informed consent. ENALEPSY was registered with ClinicalTrials.gov (NCT02332447) on 2015-01-05. The most recent Protocol and Statistical Analysis Plans are available in Appendix 3 and 4, respectively

### 2.3 Design of the ENALEPSY trial

#### – Inclusion criteria

The first patient was enrolled on 26 June 2015 and the recruitment was completed on 08 July 2020, using the following inclusion criteria: (i) Adult patient (≥ 18 years) suffering from drug-resistant focal epilepsy according to ILAE definitions and (ii) undergoing long-term video-EEG or video-stereo-electroencephalography (SEEG). Patients with history of severe heart disease or addiction to opioids, heroin, or any similar substance were excluded. Temporary administration of an immediate-release opioid agonist to manage acute pain resulted in temporary ineligibility for randomization during the 12 hours following last opioid administration whereas chronic opioid treatment was an exclusion criterion.

#### – Study drugs

NLX consisted in dihydrous hydrochloride naloxone (0.4 mg/ml) packaged in 1 ml vials. The dose of 0.4 mg corresponds to the recommended starting dose for the treatment of acute opioid overdose where the delay between intravenous injection and awakening ranges from 30 seconds to 2 minutes ^(11)^. PCB was isotonic sodium chloride prepared in 1 ml vials.

#### – Blinding and randomization

Randomization was centralized, stratified by center and balanced by block of patients to ensure 1:1 ratio between NLX and PCB. The pharmaceutical department of Hospices Civils de Lyon prepared both NLX and PCB in indistinguishable vials and numbered batches according to randomization list. The batches were then distributed to each participating centers. In case of occurrence of a FBTCS, the supervising nurse or physician took the first available batch in respect with batches numbers.

#### – Primary endpoint

The original primary endpoint was the proportion of patients with a pulse oximetry (SpO2) <90% during at least 5 seconds between 30 seconds and 5 minutes after onset of intravenous injection of the study drug in the immediate aftermath of a FBTCS^(12)^. However, in 10/2019, while the recruitment was still ongoing and before any analysis of the available data, the primary outcome was modified to the time to recovery of SpO_2_ ≥ 90% after the end of the FBTCS. This amendment of the original statistical plan, validated by the study steering committee, the independent Data and Safety Monitoring Board and the ethics committee, was justified by published data which showed that transient hypoxemia was much more frequent than previously reported^(14)^, observed in up to 86% of FBTCS ^(13)^.

#### – Secondary endpoints

The main other efficacy outcomes were: (i) SpO2 nadir; (ii) number of patients with post-ictal central apnea (PCCA); (iii) Number of patients in whom O_2_ administration was performed; (iv) Number of patients in whom cardiorespiratory rescue procedure was performed; (v) time from end of the seizure to first voluntary movement; (vi) time from end of the seizure to patient’s ability to handshake; (vii) occurrence and duration of Post-ictal Generalized EEG Suppression (PGES), as defined previously^(15)^.

The main safety outcomes were: (i) number of patients who have a second FBTCS within 120 minutes after the intravenous injection; (ii) Report of adverse events observed throughout the study.

#### – Study conduct

All included patient benefited from continuous monitoring of SpO2 and were equipped with a peripheral venous catheter throughout the video-EEG/SEEG monitoring.

Patients were continuously supervised by one to two specialized nurses during daytime in all 15 participating centres, while only six (40%) ensured 24/24 supervision. Non supervised FBTCS did not result in randomization and the patient remained eligible for randomization if another FBTCS later occured during monitoring. The management of the presurgical evaluation, including antiseizure drugs withdrawal, were not modified by the participation to the study.

Upon the occurrence of a FBTCS, treatment (NLX or PCB) was administered by the supervising nurse or physician, using direct intravenous injection within the 2 minutes following the end of the FBTCS. Importantly, treatment was never administrated before the clinical cessation of the clonic jerks. In the immediate post-ictal period, SpO2, heart and respiratory rates were carefully monitored by the supervising nurse or physician. Considering the objectives of the study, oxygen administration using high concentration breathing mask (15 l/min) was only recommended in patients who showed Sp02 < 85% for more than two minutes after the injection of the study drug. Investagors were also asked to specifically assess the ability to meet one single verbal command (handshake) in order to evaluate recovery of consciousness.

All digital data (video, EEG, SpO2) were centralized at Hospices Civils de Lyon.

#### – Analysis of seizure-related clinical data

The coordinator investigator (SR) reviewed all raw video-EEG data to: (i) confirm the times of seizure onset, onset of FBTCS, end of FBTCS and treatment injection; (ii) analyze the FBTCS semiology using the classification previously published ^(13)^; (iii) determine the end of post-ictal immobility, defined as the first voluntary movement observed on the video. Isolated myoclonic jerks that might occur after the end of a FBTCS or modifications of the patient’s position by the nurses (i.e lateral safety position) while the patient was still in post-ictal coma were not considered as voluntary movements.

#### – Analysis of SpO2 data

The evolution of the SpO2 during the 3 minutes preceding seizure onset, the seizure itself, and the 10 minutes following the end of the FBTCS, was assessed blindly to other data both by the coordinator investigator (SR) and the statisticians (FC and PRo) using a complementary approach. In both cases, postictal hypoxemia was defined as SpO2 <90% during at least 5 seconds. As previously reported^(13)^, the SpO2 signal synchronized with the video was analyzed to determine the period(s) during which SpO2 were not informative because of patient’s movements or because the oximetry sensor was not appropriately positioned on the patient’s finger. In parallel, the statistician team extracted the SpO2 signal from the video-EEG files and applied an objective analysis of SpO2 values using a specific in-house detection algorithm which allowed automatic detection and quantification of SpO2 variations. Data collected with the two approaches were then combined. When the duration of the post-ictal hypoxemia differed by ± 5 seconds between the two approaches, the value identified with the automatic detection was preferred. In case of larger differences, data were reanalyzed by both the coordinating investigator and the statisticians (CM and FC) to reach a consensus. In all analyses, the SpO2 nadir was to the lowest available value of the SpO2.

#### – Analysis of post-ictal central apnea

Two investigators (VR and SR) independently searched for the presence of post-ictal apnea using EEG breathing artifact and visual inspection of thoracoabdominal excursions on the video from the end of the clonic phase up to the patient’s first voluntary movements ^(16)^. In case of disagreement, the investigators discussed to reach consensus. Post-ictal central apnea (PCCA) was defined as lack of breath for a period > 5 seconds. PCCA was considered immediate if no breath was taken during the 5 seconds following seizure end ^(17)^.

#### – Sample size

We based our hypothesis on our previously reported SpO2 findings following FBTCS with and without O_2_ therapy, showing that the proportion of patients who recovered SpO2 ≥ 90% at 30, 60 and 120 seconds post-FBCTS was 17%, 52% and 88% without O_2_ administration, and 48%, 79% and 92% with O_2_ administration.^(13)^. We hypothesized that the size of the NLX effect to reduce the recovery time to SpO2 >90% would be similar to that of oxygen therapy, with a hazard ratio of 2.4. Accordingly, using a log rank test with a two-sided alternative hypothesis (significance level of 5%), 40 events needed to be observed to reject the null hypothesis in 80% of cases.

### 2.3 Historical control (HC)

#### – Patients’ and FBTCS selection

For HC, we used the data from patients who participated in the REPO_2_MSE study and whose SpO2 data following FBTCS were previously published ^(13)^. According to their comparable inclusion criteria, virtually all patients included in the REPO_2_MSE study would have been eligible for inclusion in the ENALEPSY study. We then reviewed the video of the 107 available FBCTS to determine those which would have fulfilled criteria for randomization in the ENALEPSY trial, with a specific attention to the lack of early administration of oxygen and quality of video to assess duration of post-ictal immobility and coma (figure 1). When several FBCTS from the same patient fulfilled these criteria, only the first FBCTS recorded during the hospital stay was included in the HC group. Data extraction for HC was performed using the same methodology as that described above for patients recruited in the ENALEPSY trial.

**Figure 1:**
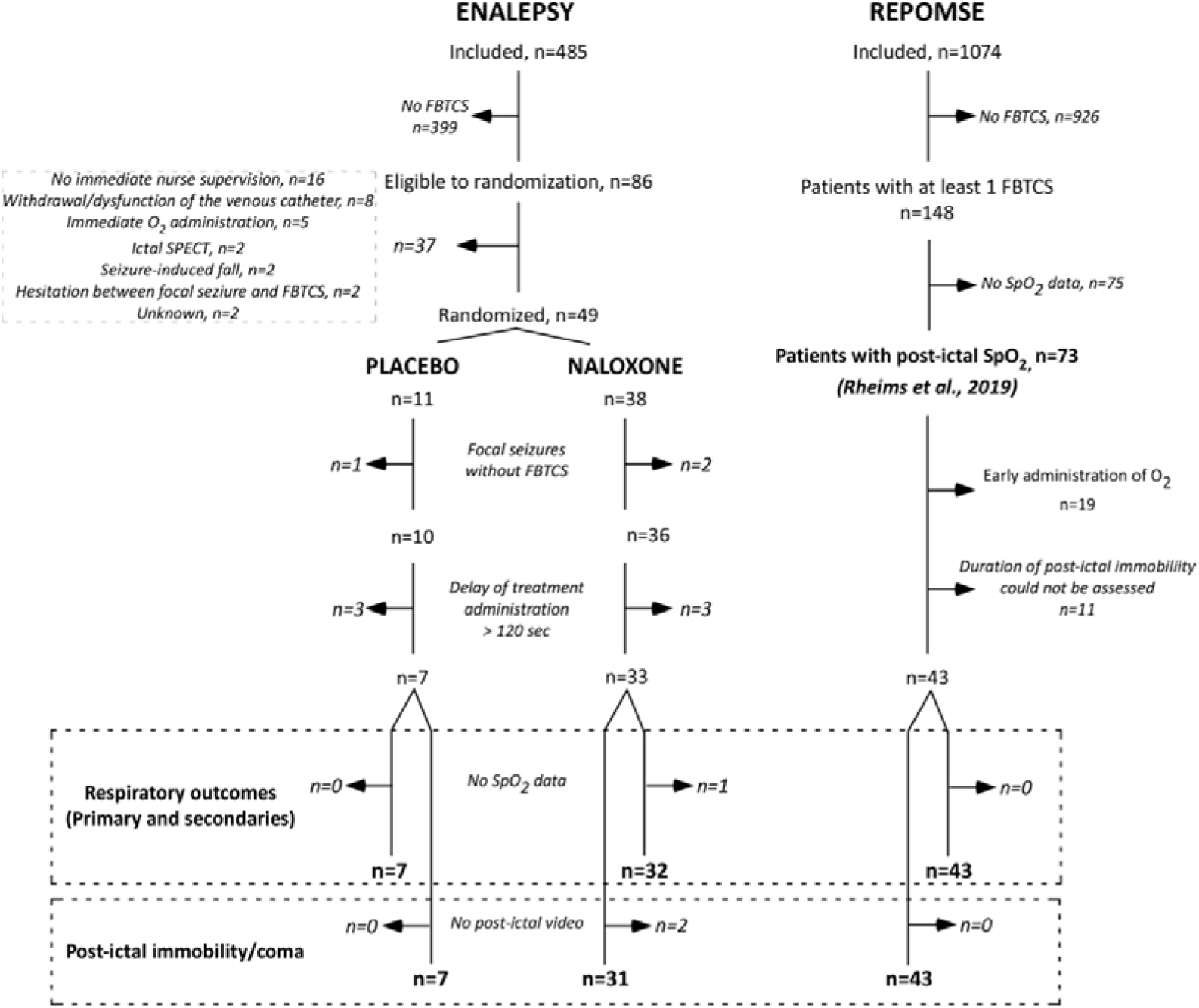
Flowchart of the study.

**Figure 2:**
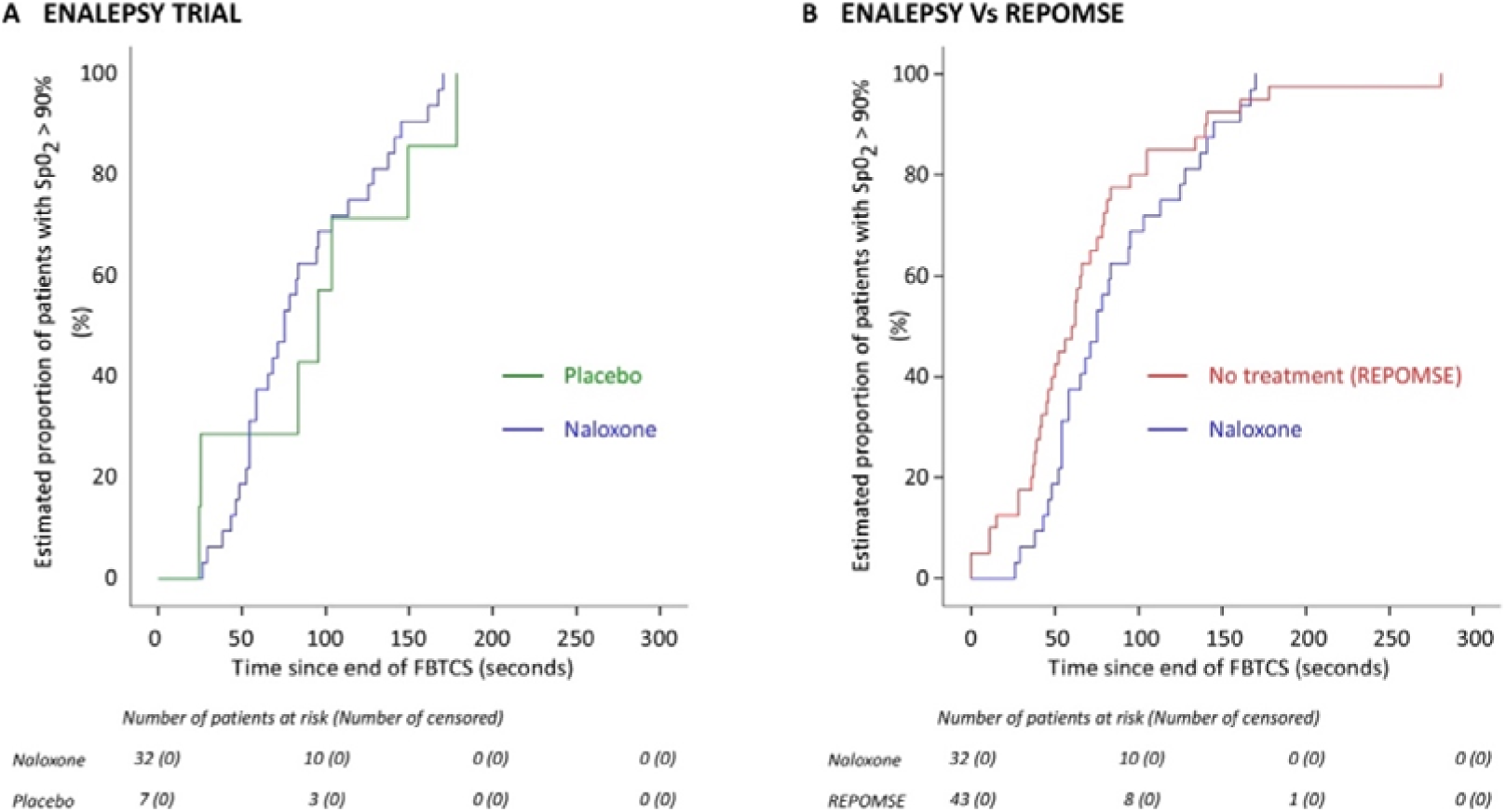
Time to recovery of SpO2 ≥ 90% from the end of the seizure to 5 min post-ictal.

### 2.4 Statistical analyses

#### – Populations

Because of the major protocol violation related to the mismanagement from the pharmaceutical department, intention to treat analysis could not be performed. All efficacy analyses were therefore performed in the per-protocol population (ENALEPSY PP) defined as all randomized patients with FBTCS in whom the study drug was administered > 120 seconds after the end of the seizure. The safety population corresponded to all randomized patients.

#### – Primary endpoint

The Kaplan-Meier estimator was used to estimate the evolution of the probability of recovery of SpO2 ≥ 90% between the end of the FBTCS and 5 minutes post-ictal. An accelerated failure time model considering a Weibull distribution was used to estimate hazard ratio (HR) of NLX vs PCB and NLX vs HC effects ^(18)^.

#### – Secondary endpoints

SpO2 nadir and proportion of FBTCS associated with PCCA or PGES were compared between groups using Mann-Whitney or Fischer test. Time to first voluntary movement and to patient’s ability to handshake was analysed from the end of the seizure to 5 min post-ictal with the same survival analysis as for the primary endpoint.

In all analyses, treatment effect estimates are presented with 95% confidence intervals (CIs). Two-sided P values less than 0.05 were considered significant.. All analyses were performed using SAS ®version 9.4 or further (SAS Institute Inc., Cary, NC, USA.) or R version 4.0.2 or further (R Core Team (2012).

## 3. RESULTS

### 3.1 Populations

#### – ENALEPSY trial

The flowchart of the study is detailed in figure 1. Among the 485 patients included in ENALEPSY, 86 had at least one FBTCS and were eligible for randomization. However, mainly because of lack of immediate nurse supervision or lack of available catheter, only 49 patients were eventually randomized. No difference was observed between eligible patients who were eventually randomized and those who were not (supplemental table 1). Among the 49 randomized patients, 11 received PCB and 38 NLX. After exclusion of patients with protocol violations (i.e delay of treatment administration > 120 sec), 40 patients (7 PCB and 33 NLX) were included in the per protocol population (ENALEPSY PP).

#### – Historical control

After exclusion of patients who had received oxygen during all their FBTCS (n=19) and those in whom post-ictal coma could not be assessed (n=11), 43 of the 73 patients from the REPO_2_MSE cohort ^(13)^ were included in the HC group.

### 3.2 Patients’ and seizures’ characteristics

The detailed characteristics of the patients and the FBTCS are presented in tables 1 and 2. Patients’ characteristics were comparable between the ENALEPSY PP study and the HC group. The two exceptions were the FBTCS frequency over the last 12months, with greater proportion of patients with ≥ 1 FBTCS / month in ENALEPSY PP than in the HC group. The type of FBTCS was also comparable across groups, with 51.2%, 16.3% and 32.6% of FBTCS type 1, 2 and 3, respectively, in the HC group and 57.9%, 13.2% and 28.9% in ENALEPSY PP. Nurse intervention during the course of the seizure and in the post-ictal period was observed in all FBTCS in ENALEPSY PP and in all but three (93%) in the HC group. For these later, nurse intervention during the FBTCS was not maintained in the post-ictal period in one whereas there was no intervention in the two others. A total of 22 FBTCS (51.2%) occurred during sleep in the HC group, versus 11 (27.5%) in ENALEPSY PP. A transient loss of SpO2 signal was observed in 26 (66%) FBTCS in ENALEPSY PP and in 28 (65%) in the HC group, with a mean duration of the period without informative SpO2 ± SD of 50.5 ± 48.3 and 18.1 ± 21.2 seconds, respectively.

**Table 1:** Characteristics of patients included in ENALEPSY PP and in the HC group.

|  | ENALEPSY Study (PP population) |  |  | **REPOMSE Study (HC group)**<br>**(N=43)** |
| --- | --- | --- | --- | --- |
|  | *All patients (n=40)* | *Placebo (N=7)* | *Naloxone (N=33)* |  |
| **Age, mean (sd)** | 35.6 (11.6) | 34.71 (18.4) | 35.76 (9.95) | 33.98 (10) |
| **Gender, n (%)** |  |  |  |  |
| *Female* | 18 (45.0%) | 4 (57.1%) | 14 (42.4%) | 20 (46.5%) |
| **Epilepsy duration, mean (sd)** | 16.1 (9.5) | 16.71 (14.27) | 15.94 (8.42) | 16.82 (10.16) |
| **Seizure frequency, n (%)** |  |  |  |  |
| $\geq 1$ / day | 5 (12.5%) | 2 (33.3%) | 3 (9.1%) | 4 (9.3%) |
| $< 1/d$ but $\geq 1/wk$ | 15 (37.5%) | 2 (33.3%) | 13 (39.4%) | 13 (30.2%) |
| $< 1/wk$ | 20 (50%) | 3 (42.8%) | 17 (51.5%) | 25 (58.1%) |
| Unknown | 0 (0%) | 0 (0%) | 0 (0%) | 1 (2.3%) |
| **FBTCS frequency, n (%)** |  |  |  |  |
| $\geq 1/mo$ | 15 (37.5%) | 1 (14.3%) | 14 (42.4%) | 5 (11.6%) |
| $< 1/mo$ but $\geq 1/y$ | 6 (15%) | 2 (33.3%) | 4 (12.1%) | 15 (34.9%) |
| $< 1/y$ | 15 (37.5%) | 4 (57.1%) | 11 (33.3%) | 19 (44.2%) |
| None | 0 (0%) | 0 (0%) | 0 (0%) | 0 (%) |
| Unknown | 4 (10%) | 0 (0%) | 4 (12.1%) | 4 (9.3%) |
| **Past epilepsy surgery, n (%)** | 5 (12.5%) | 0 (0%) | 5 (15.2%) | 3 (7%) |
| **Sleep apnea syndrome, n (%)** | 1 (2.5%) | 0 (0%) | 1 (3.0%) | 1 (2.3%) |
| **Treatment by VNS, n (%)** | 1 (2.5%) | 0 (0%) | 1 (3.0%) | 3 (7%) |
| **Number of ASM, n (%)** |  |  |  |  |
| 0 | 0 (0%) | 0 (0%) | 0 (0%) | 2 (4.7%) |
| 1 | 7 (21.9%) | 2 (33.3%) | 5 (19.2%) | 5 (11.6%) |
| 2 | 15 (46.9%) | 2 (33.3%) | 13 (50.0%) | 22 (51.2%) |
| $\geq 3$ | 10 (31.2%) | 2 (33.3%) | 8 (30.8%) | 14 (32.6%) |
| **Treatment with SSRI, n (%)** | 0 (0%) | 0 (0%) | 0 (0%) | 3 (7.1%) |
| **Seizure focus, n (%)** |  |  |  |  |
| *Temporal* | 23 (57.5%) | 6 (85.7%) | 17 (51.5%) | 21 (48.9%) |
| *Extra-Temporal* | 16 (40%) | 1 (14.3%) | 15 (45.5%) | 19 (44.2%) |
| Unknown | 1 (2.5%) | 0 (0%) | 1 (3.0%) | 3 (6.9%) |

VNS: vagal nerve stimulation; ASM: antiseizure medication; SSRI: selective serotonin recapture inhibitor

**Table 2:** Characteristics of FBTCS.

|  | ENALEPSY Study (PP population) |  |  | **REPOMSE Study (HC group)**<br>(N=43) |
| --- | --- | --- | --- | --- |
|  | *All patients*<br>(n=40) | *Placebo*<br>(N=7) | *Naloxone*<br>(N=33) |  |
| **Recordings modalities** |  |  |  |  |
| *SEEG* | 7 (17.5%) | 1 (14.3%) | 6 (18.8%) | 0 (0%) |
| **Seizure type\*** |  |  |  |  |
| *I* | 22 (57.9%) | 2 (33.3%) | 20 (62.5%) | 22 (51.2%) |
| *II* | 5 (13.2%) | 0 (0%) | 5 (15.6%) | 7 (16.3%) |
| *III* | 11 (28.9%) | 4 (66.7%) | 7 (21.9%) | 14 (32.6%) |
| **Duration of tonic phase, seconds, mean (sd)\*** | 13.2 (7.6) | 17 (8.02) | 12.5 (7.47) | 12.3 (7.28) |
| **Duration of tonic-clonic phase, seconds, mean (sd)\*** | 55.5 (13.4) | 64.33 (14.31) | 53.84 (12.8) | 51.74 (12.39) |
| **State of wakefulness** |  |  |  |  |
| *Asleep* | 11 (27.5%) | 2 (28.6%) | 9 (27.3%) | 22 (51.2%) |
| **Administration of O2** |  |  |  |  |
| *Early administration* | 2 (5%) | 0 (0%) | 2 (6.1%) | 0 (0%) |
| *Delayed administration* | 1 (2.5%) | 0 (0%) | 1 (3.0%) | 5 (11.6%) |
| **Nurse intervention during seizure** | 40 (100%) | 7 (100%) | 33 (100%) | 40 (93%) |

\* data available in 32 patients in the Naloxone group and 6 in PCB group

The mean ± SD time from termination of FBTCS to study drug or placebo injection was 44.2±32.2 seconds (range 0-118), without difference between the NLX group (44.9±32.0 seconds) and the PCB group (44.4±35.6 seconds). However, SpO2 was already ≥ 90% in 9 patients when the treatment was administered, including 8 NLX (24%) and 1 PCB (14%)).

### 3.3 Efficacy of NLX

#### – Respiratory endpoints

The mean time ± SD from the termination of FBTCS to recovery of SpO2 ≥ 90% was 85.5±41.7 seconds with NLX, 93.9±57.7 seconds with PCB and 70.2±54.1 seconds in the HC group (table 3). Administration of NLX did not accelerate recovery of SpO2 ≥ 90% in comparison with PCB (HR (95%CI): 1.33 (0.59 – 3.01), p=0.497) or with HC (HR (95%CI): 0.88 (0.55 – 1.41), p=0.607). The results remained similar when time from treatment administration to recovery of SpO2 ≥ 90% was considered (71.1±33.2 seconds with NLX and 68.5±42.0 seconds with PCB). Similarly, the SpO2 nadir was similar in NLX, PCB and HC groups (Table 3). Occurrence of PCCA was similar across groups (29% in NLX, 14.3% in PCB and 30.2% in HC; table 3), including when only delayed PCCA were considered (12.9% in NLX, 14.3% in PCB and 16.3.2% in HC). Similarly, the mean ± SD duration of PCCA did not differ across groups (Table 3). No cardio-respiratory rescue procedure was required in any of the randomized patients.

**Table 3:**
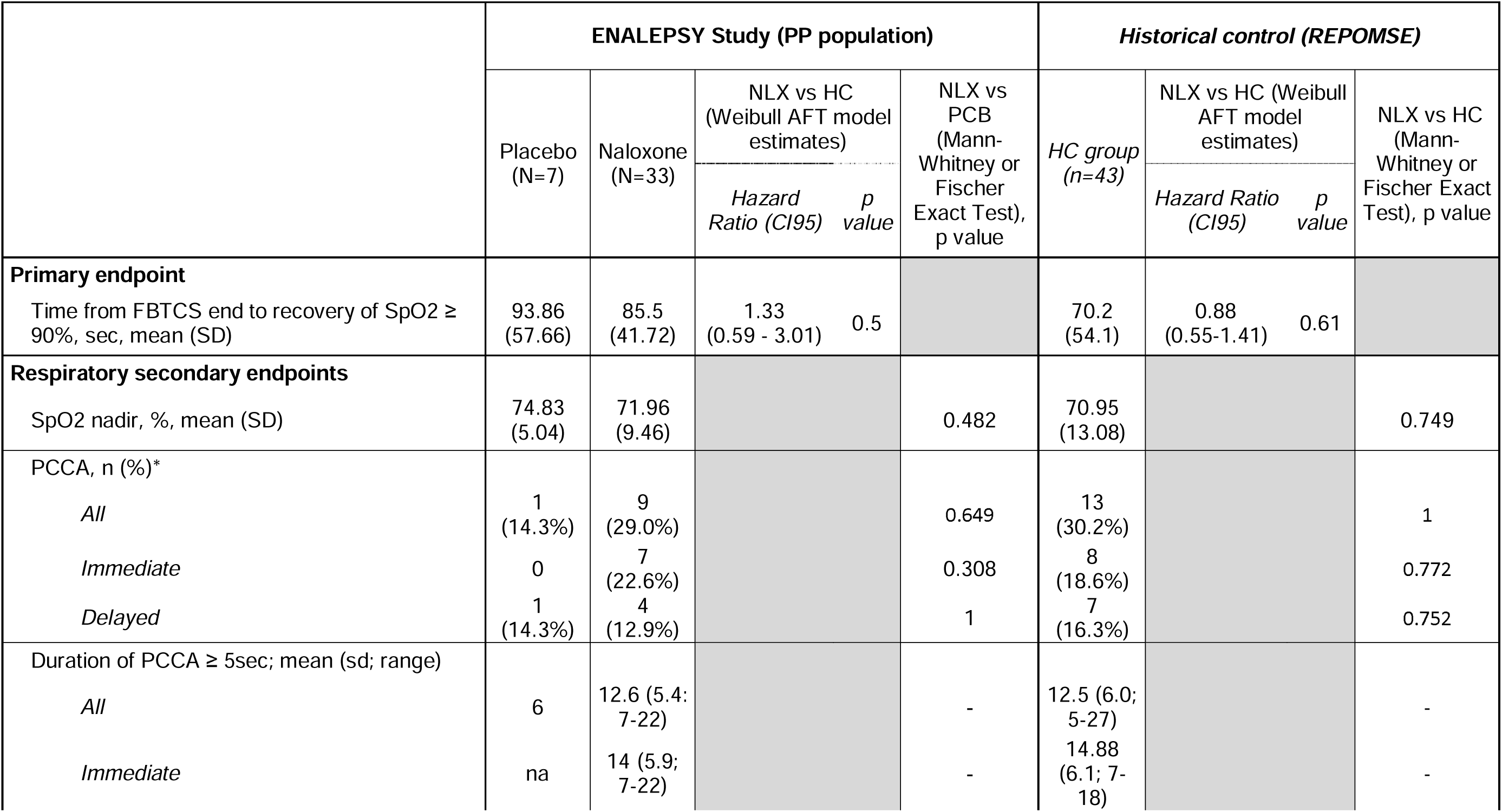
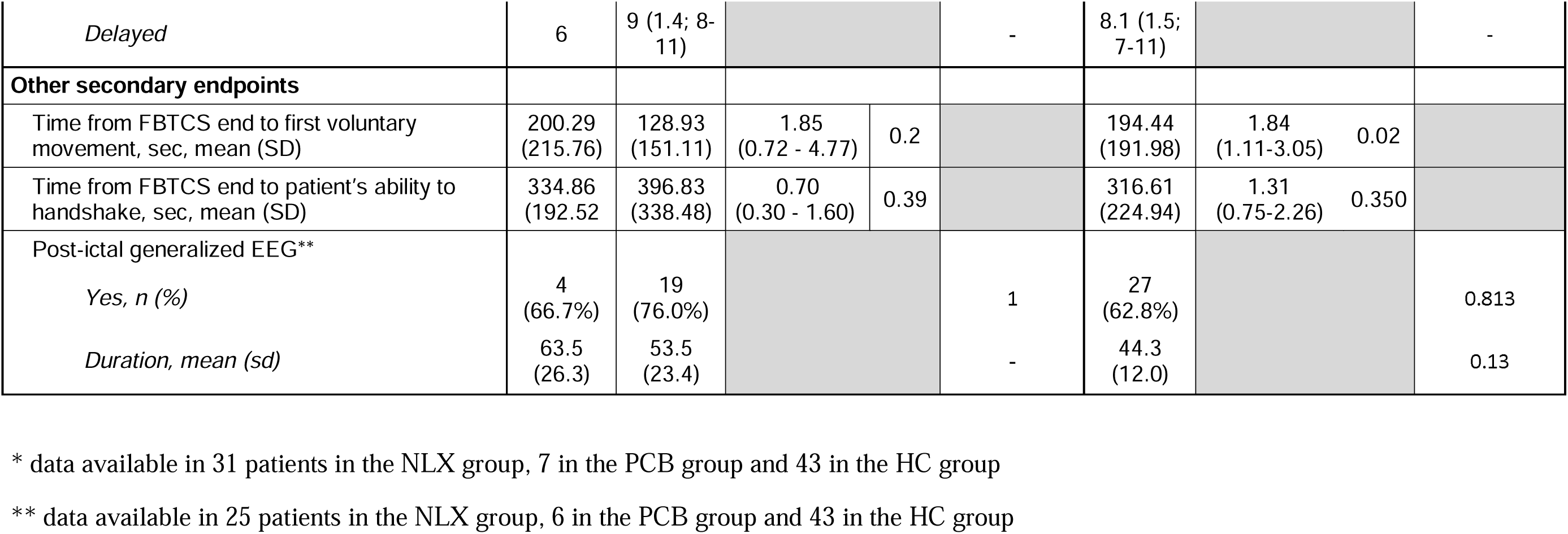
Primary and secondary endpoints.

|  | ENALEPSY Study (PP population) |  |  |  | *Historical control (REPOMSE)* |  |  |  |  |
| --- | --- | --- | --- | --- | --- | --- | --- | --- | --- |
|  | Placebo<br>(N=7) | Naloxone<br>(N=33) | NLX vs HC<br>(Weibull AFT model<br>estimates) |  | NLX vs<br>PCB<br>(Mann-Whitney or<br>Fischer<br>Exact Test),<br>p value | *HC group*<br>(n=43) | NLX vs HC (Weibull<br>AFT model<br>estimates) |  | NLX vs HC<br>(Mann-Whitney or<br>Fischer Exact<br>Test), p value |
|  |  |  | *Hazard*<br>*Ratio (CI95)* | *p*<br>*value* |  |  | *Hazard Ratio*<br>*(CI95)* | *p*<br>*value* |  |
| **Primary endpoint** |  |  |  |  |  |  |  |  |  |
| Time from FBTCS end to recovery of SpO2 ≥ 90%, sec, mean (SD) | 93.86<br>(57.66) | 85.5<br>(41.72) | 1.33<br>(0.59 - 3.01) | 0.5 |  | 70.2<br>(54.1) | 0.88<br>(0.55-1.41) | 0.61 |  |
| **Respiratory secondary endpoints** |  |  |  |  |  |  |  |  |  |
| SpO2 nadir, %, mean (SD) | 74.83<br>(5.04) | 71.96<br>(9.46) |  |  | 0.482 | 70.95<br>(13.08) |  |  | 0.749 |
| PCCA, n (%)* |  |  |  |  |  |  |  |  |  |
| *All* | 1<br>(14.3%) | 9<br>(29.0%) |  |  | 0.649 | 13<br>(30.2%) |  |  | 1 |
| *Immediate* | 0 | 7<br>(22.6%) |  |  | 0.308 | 8<br>(18.6%) |  |  | 0.772 |
| *Delayed* | 1<br>(14.3%) | 4<br>(12.9%) |  |  | 1 | 7<br>(16.3%) |  |  | 0.752 |
| Duration of PCCA ≥ 5sec; mean (sd; range) |  |  |  |  |  |  |  |  |  |
| *All* | 6 | 12.6 (5.4;<br>7-22) |  |  | - | 12.5 (6.0;<br>5-27) |  |  | - |
| *Immediate* | na | 14 (5.9;<br>7-22) |  |  | - | 14.88<br>(6.1; 7-18) |  |  | - |
| *Delayed* | 6 | 9 (1.4; 8-11) |  |  | - | 8.1 (1.5; 7-11) |  |  | - |
| **Other secondary endpoints** |  |  |  |  |  |  |  |  |  |
| Time from FBTCS end to first voluntary movement, sec, mean (SD) | 200.29<br>(215.76) | 128.93<br>(151.11) | 1.85<br>(0.72 - 4.77) | 0.2 |  | 194.44<br>(191.98) | 1.84<br>(1.11-3.05) | 0.02 |  |
| Time from FBTCS end to patient's ability to handshake, sec, mean (SD) | 334.86<br>(192.52) | 396.83<br>(338.48) | 0.70<br>(0.30 - 1.60) | 0.39 |  | 316.61<br>(224.94) | 1.31<br>(0.75-2.26) | 0.350 |  |
| Post-ictal generalized EEG** |  |  |  |  |  |  |  |  |  |
| *Yes, n (%)* | 4<br>(66.7%) | 19<br>(76.0%) |  |  | 1 | 27<br>(62.8%) |  |  | 0.813 |
| *Duration, mean (sd)* | 63.5<br>(26.3) | 53.5<br>(23.4) |  |  | - | 44.3<br>(12.0) |  |  | 0.13 |

\* data available in 31 patients in the NLX group, 7 in the PCB group and 43 in the HC group
\*\* data available in 25 patients in the NLX group, 6 in the PCB group and 43 in the HC group

#### – Other secondary endpoints

The mean time ± SD from the termination of FBTCS to the first voluntary movement was similar in the PCB (200.3±215.8 seconds) and HC groups (194.4±192 seconds) but lower in the NLX group (128.9±151.1 seconds). Accordingly, an acceleration of the time of occurrence of the first voluntary movement was observed with NLX (figure 3), which was significant when compared with no treatment (HR (95%CI); 1.84 (1.11 – 3.05), p=0.021). In contrast, the duration of post-ictal coma did not differ across groups (Table 3). PGES could be assessed in 6 PCB, 25 NLX, while not available in nine patients, seven who undergone SEEG and two whose scalp EEG electrodes became detached during the FBTCS. Neither the rate of occurrence nor the mean duration of PGES differed between groups (table 3).

**Figure 3:**
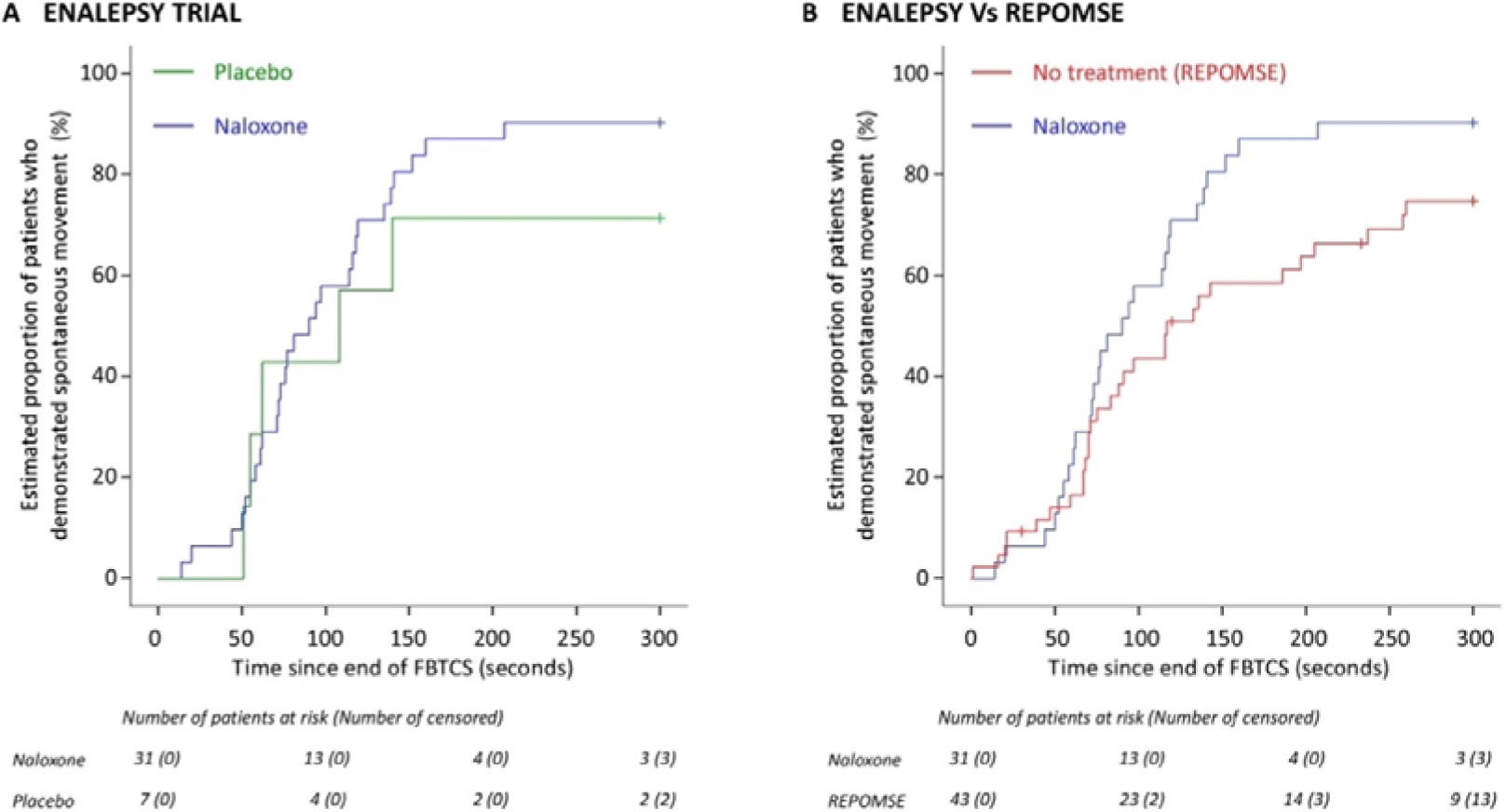
Time to first voluntary movement and to patient’s ability to handshake was analysed from the end of the seizure to 5 min post-ictal.

### 3.4 Safety of NLX

No serious adverse event was reported in the ENALEPSY study. A FBTCS recurrence within the 2 hours following treatment administration was reported in four patients who received NLX (12.1%), one patient who received PCB (14.3%) and five patients from the HC group (11.6%). Pain following the postictal coma did not differ across groups. The mean ±SD score of the visual analog scale was 1.89 ±2.95 with NLX (n=27) and 2.83 ± 3.19 with PCB (n=6).

## DISCUSSION

Results from the ENALEPY trial and comparison to historical controls suggest that post-ictal administration of NLX does not decrease the severity of post-FBTCS respiratory dysfunction, but might be associated with a reduction in the duration of post-ictal immobility.

These conclusions must be tempered by the methodological limitations of the study. Due to an error in the preparation of batches by the pharmacy resulting in a major imbalance in treatment allocation, the ENALEPSY trial could no longer be analyzed on an intention-to-treat basis. Additionally, the sample size of the group that eventually received PCB was too small to ensure sufficient statistical power. For these reasons, we changed the design of this double-blind RCT into an exploratory study, whereby patients who received NLX were compared to an untreated group of patients whose data were collected in the previously reported REPO_2_MSE cohort ^(13)^. Importantly, patients’ and FBTCS characteristics were similar between the ENALEPSY population and the control HC group, whose data were collected from the same EMUs. Accordingly, the primary and secondary endpoints also proved similar between the ENALEPSY PCB group and the HC group, particularly the duration of post-ictal immobility.

Post-ictal respiratory dysfunction was primarily evaluated by monitoring the occurrence of peri-ictal hypoxemia using SpO2 and video analysis of thoracoabdominal excursions to detect post-ictal central apneas. Although this video-based approach was used previously ^(5; 16)^, its precision remains lower than inductance plethysmography.

The lack of observable impact of NLX on hypoxemia could be due to a too low dose of NLX. In case of morphine overdose, the initial NLX dose is 0.4 mg, as in our study, but dose escalation up to 15 mg can then be performed based on clinical response, with NLX efficacy defined by increase in respiratory rate^(11)^. A higher dose of NLX might thus have shown efficacy on our primary endpoint. Yet, the effect of NLX on the duration of post-ictal immobility suggests that 0.4 mg was sufficient to produce a biological effect. According to its pharmacokinetics^(19)^, NLX effect is expected between 30 seconds to 2 minutes after intravenous administration, and its apparent duration of action is 20 to 90 minutes^(11)^. In this context, a NLX effect could not be expected for endpoints evaluated within the first 30 seconds after the end of the clonic phase, including immediate PCCA and PGES occurrence. The delay from termination of FBTCS to injection of NLX might have also reduced the probability to detect the impact of NLX on other respiratory endpoints. However, the mean time to recovery of SpO2 ≥ 90% in the 14 patients (42.4%) who received NLX within the first 30 seconds (85±38 seconds) was comparable to that of patients who received NLX after a longer delay (85.9±45.7 seconds), suggesting that the impact of NLX on SpO2 recovery did not depend on this delay.

Although current data argue for a predominant role of central apnea in the pathophysiology of SUDEP, the role of post-ictal coma severity must also be considered ^(20)^. The duration of post-ictal immobility has been reported to be associated with the risk of PGES occurrence and the severity of respiratory dysfunction^(21)^. Moreover, it can be speculated that a longer duration of post-ictal immobility might increase the risk of prolonged post-ictal prone position and associated obstructive respiratory disorder. This issue might is particularly important for seizures occurring during sleep, which represent an independent risk factor of SUDEP. Several mechanisms might participate to this association, including alteration of arousal reactivity in drug-resistant epilepsy ^(22)^, especially asphyxia-induced arousal ^(23)^, and increased severity of post-ictal coma ^(24)^. In our study, sleep-onset FBTCS were more frequent in the HC group (51.2%) than in ENALEPSY PP (27.5%), reflecting more frequent randomizations during daytime. Some studies thus suggested that PGES are more frequent after sleep than day-time FBTCS ^(25)^, though this observation remains debated ^(15)^. On the other hand, neither post-FBTCS respiratory dysfunction ^(13;16; 25)^ nor the duration of post-ictal immobility ^(25)^ was reported to differ between sleep-onset and wake-onset FBTCS.

Because of the role of endogenous opioids in post-ictal seizure inhibition, administration of NLX might have increased the risk of an early recurrence of FBTCS. Our data showed that this was not the case. However, we cannot exclude that higher doses of NLX might have a different safety profile.

In conclusion, although our study failed to meet its primary endpoint, with no efficacy of NLX on post-FBTCS respiratory dysfunction, it suggests a potential role for opiate antagonists in mitigating other post-ictal disorders that might contribute to FBCTS-induced SUDEP. In the event that this acute effect of NLX would be confirmed, another long-acting opioid antagonist, naltrexone, could be tested as a chronic therapy to reduce the risk of SUDEP in persons with frequent FBCTS. Naltrexone has proved a safe drug in very large cohorts of patients with chronic alcoholism who are often at risk of breakthrough generalized convulsive seizures, without reports suggesting an increased risk of such seizure, in line with the lack of increased risk of seizure recurrence with NLX in our study.

## Supporting information

CONSORT Checklist

ENALEPSY Study Group

Full protocol of the ENALEPSY Study

ENALEPSY Statistical Plan

## Data Availability

All data produced in the present study are available upon reasonable request to the authors

## Acknowledgments

The ENALEPSY study was funded by the French Ministry of Health (PHRC National 2013). The REPO_2_MSE study was funded by the French Ministry of Health (PHRC National 2009). Vincent Rossero is funded by ERANET-NEURON – Agence Nationale de la Recherche, Grant/ Award Number: ANR-21-NEU2-0006 Sylvain Rheims is funded by HORIZON-ERC-Award Number: ERC-2023-COG-101125118

## Ethical Publication Statement

We confirm that we have read the Journal’s position on issues involved in ethical publication and affirm that this report is consistent with those guidelines.

---

Supplemental Table S1.

**Supplemental Table S1:**
|  | All patients<br>(N=485) | Eligible for randomization |  |  |
| --- | --- | --- | --- | --- |
|  |  | All<br>(n=86) | Not<br>randomized<br>(N=37) | Randomized<br>(N=49) |
| **Age, mean (sd)** | 36.5 (11.5) | 33.5 (11.4) | 31.73 (11.61) | 34.82 (11.11) |
| **Gender, n (%)** |  |  |  |  |
| *Female* | 261 (53.8%) | 47 (54.7%) | 23 (62.2%) | 24 (49.0%) |
| **Epilepsy duration, mean (sd)** | 17.9 (11.5) | 15.9 (10.4) | 15.64 (11.97) | 16.17 (9.25) |
| **Body Mass Index, mean (sd)** | 26.7 (30.6) | 24.4 (6.0) | 25.71 (6.5) | 23.5 (5.52) |
| **Seizure frequency, n (%)** |  |  |  |  |
| $\geq 1$ / day | 68 (14%) | 8 (9.3%) | 2 (5.4%) | 6 (12.2%) |
| $< 1/d$ but $\geq 1/wk$ | 110 (22.7) | 31 (36.0%) | 10 (27.0) | 21 (42.9%) |
| $< 1/wk$ | 291 (60%) | 46 (53.5%) | 25 (67.6%) | 21 (42.9%) |
| Unknown | 16 (3.3%) | 1 (1.2%) | 0 (0%) | 1 (2.0%) |
| **FBTCS frequency, n (%)** |  |  |  |  |
| $\geq 1/mo$ | 112 (23.1%) | 35 (40.7%) | 16 (43.2%) | 19 (38.8%) |
| $< 1/mo$ but $\geq 1/y$ | 54 (11.1%) | 14 (16.3%) | 8 (21.6%) | 6 (12.2%) |
| $< 1/y$ | 61 (12.6%) | 20 (23.3%) | 4 (10.8%) | 16 (32.7%) |
| None | 237 (48.9%) | 11 (12.8%) | 9 (24.3%) | 2 (4.1%) |
| Unknown | 21 (4.3%) | 6 (7.0%) | 0 (0%) | 6 (12.2%) |
| **Past epilepsy surgery, n (%)** | 34 (7%) | 7 (8.1) | 1 (2.7%) | 6 (12.2%) |
| **Treatment by VNS, n (%)** | 19 (3.9%) | 1 (1.2%) | 0 (0%) | 1 (2.0%) |
| **Number of ASM, n (%)** |  |  |  |  |
| 0 | 2 (0.4%) | 1 (1.2) | 1 (2.7%) | 0 (0%) |
| 1 | 56 (11.5%) | 11 (12.8) | 4 (10.8%) | 7 (14.3%) |
| 2 | 196 (40.4%) | 39 (45.3) | 19 (51.4%) | 20 (40.8%) |
| $\geq 3$ | 231 (47.6%) | 35 (40.7%) | 13 (35.1%) | 22 (44.9%) |
| **Treatment with SSRI, n (%)** | 0 (0%) | 0 (0%) | 0 (0%) | 0 (0%) |
| **Type of epilepsy, n (%)** |  |  |  |  |
| *Focal epilepsy* | 465 (95.9%) | 85 (98.8%) | 36 (97.3%) | 49 (100%) |
| *Idiopathic generalized epilepsy* | 4 (0.8%) | 1 (1.2) | 1 (2.7%) | 0 (0%) |
| *PNES* | 16 (3.3%) | 0 (0%) | 0 (0%) | 0 (0%) |
| **Seizure focus of patients with focal epilepsy, n (%)** |  |  |  |  |
| *Temporal* | 236 (50.8%) | 45 (52.9%) | 19 (52.8%) | 26 (53.1%) |
| *Extra-Temporal* | 100 (21.5%) | 25 (29.4%) | 11 (30.6%) | 8 (16.3%) |
| *Multilobar* | 36 (7.7%) | 11 (12.9%) | 3 (8.3%) | 11 (12.8%) |
| Unknown | 93 (20%) | 4 (4.7%) | 3 (8.3%) | 4 (4.7%) |
| **SEEG** | 75 (16.0%) | 14 (16.3) | 5 (13.5%) | 9 (18.4%) |

