## Supplementary material for "Efficacy of Naloxone in reducing hypoxemia and duration of immobility following focal to bilateral tonic-clonic seizures": ENALEPSY Study Group

**List of investigators of the ENALEPSY study**

Bordeaux: Drs V. Michel, M. De Montaudouin

Dijon: Dr M. Lemesle Martin

Grenoble: Pr P. Kahane, Drs L. Minotti, L. Vercueil, AS Jobst

La Teppe: Dr J. Petit, Dr D. Tourniaire, Dr V. Eid, Dr P. Latour

Lille: Pr P. Derambure, Dr W. Szurhaj,

Lyon: Pr S. Rheims, Dr H. Catenoix, Dr N. Andre-Obadia, Dr J. Isnard, Dr A. Montavont, Dr S. Boulogne

Marseille: Prs F. Bartolomei et. A.Trébuchon ; Drs A. Mc Gonigal, S. Aubert, S Lagarde

Montpellier: Drs A. Crespel, P. Gelisse, B. Mercedes

Nancy: Pr L. Maillard, Pr L. Tyvaert Dr JP Vignal

Paris Pitié-Salpêtrière: Prs V. Navarro, S. Dupont, Dr C. Adam, Dr V. Frazzini, Dr V-H Nguyen-Michel, Dr M Damiano, Dr V Lambrecq

Rennes: Drs A. Biraben, A. Nica

Saint-Etienne: Dr P. Convers, Dr L. Mazzola

Strasbourg: Pr E. Hirsch, Dr MP. Valenti, Dr J. Scholly, Dr C. Behr

Toulouse: Drs L. Valton, M. Denuelle, J. Curot

Tours: Dr J. Biberon
