## Supplementary material for "Efficacy of Naloxone in reducing hypoxemia and duration of immobility following focal to bilateral tonic-clonic seizures": Full protocol of the ENALEPSY Study

### ENALEPSIE

#### EFFICACY OF NALOXONE IN REDUCING POSTICTAL CENTRAL RESPIRATORY DYSFUNCTION IN PATIENTS WITH EPILEPSY.

Version N°10 - Date: 23/02/2021
N° EudraCT: 2014-003003-30

##### COORDINATOR INVESTIGATOR

**Pr Sylvain RHEIMS**
Service de Neurologie Fonctionnelle et d'Epileptologie et Institut des Epilepsies
Hôpital Neurologique
Hospices Civils de Lyon
59, boulevard Pinel
F - 69677 Bron Cedex
Tel.: + 33 4 72 35 70 44
Fax: + 33 4 72 11 80 39


Sylvain Rheims le 23 février 2021

##### METHODOLOGY,

**Unité Recherche Clinique**
**Pôle Information Médicale, Evaluation et Recherche**
Hospices Civils de Lyon
162, avenue Lacassagne
F - 69424 Lyon Cedex 03

**Pr François CHAPUIS**
Tel.: + 33 4 72 11 51 68 / Fax : + 33 4 72 11 57 11


**Dr Géraldine SAMSON**
Tel.: + 33 4 72 11 57 74 / Fax: + 33 4 72 11 57 11


#### COORDINATION AND DATA ANALYSIS CENTRE

**Service de Biostatistique-Bioinformatique des HCL**
Equipe Biostatistique Sante UMR 5558 CNRS UCBL
162 Avenue Lacassagne
69424 Lyon Cedex 03 FRANCE

**Pr ROY Pascal**

**CATHERINE MERCIER**

#### SPONSOR

**Hospices Civils de Lyon**
3, quai des Célestins
F - 69229 Lyon Cedex 02

*Person authorized to sign the protocol and the protocol amendment(s) for the sponsor:*

**Mme DION Armelle**
Délégation à la Recherche Clinique et à l'Innovation – Hospices Civils de Lyon

*Responsible for all trial-site related medical decisions:*

**Dr Valérie PLATTNER**
Délégation à la Recherche Clinique et à l'Innovation – Hospices Civils de Lyon
Tél. : + 33 4 72 40 68 40 / Fax : + 33 4 72 11 51 90

*Responsible of safety and pharmacovigilance team:*

**Marina NGUON**
Pharmacien référent
Délégation à la Recherche Clinique et à l'Innovation – Hospices Civils de Lyon
Tél : + 33 4 72 40 68 26 / Fax : + 33 4 72 11 51 90

#### STUDY STERRING COMMITTEE

Pr Sylvain RHEIMS, Service de Neurologie Fonctionnelle et d'Epileptologie, Hospices Civils de Lyon
Pr Philippe RYVLIN, Service de Neurologie Fonctionnelle et d'Epileptologie, Hospices Civils de Lyon
Pr Fabrice BARTOLOMEI, Centre Hospitalier Universitaire de la Timone, Marseille
Pr François CHAPUIS, Unité Recherche Clinique, Pôle Information Médicale, Evaluation et Recherche, Hospices Civils de Lyon, Lyon.

#### LIST OF INVESTIGATORS

| Last Name | First Name | City | Country | Hospital | email | Phone | Speciality |
|---|---|---|---|---|---|---|---|
| BARTOLOMEI | Fabrice | Marseille | France | Centre Hospitalier Universitaire de la Timone | | 0491385833 | Neurology |
| DERAMBURE | Philippe | Lille | France | Centre Hospitalier Régional et Universitaire Roger Salengro | | 0320446373 | Neurology |
| HIRSCH | Edouard | Strasbourg | France | Hôpitaux Universitaires de Strasbourg | | 0388116768 | Neurology |
| MAILLARD | Louis | Nancy | France | Centre Hospitalier Universitaire de Nancy | | 0383851686 | Neurology |
| MICHEL | Véronique | Bordeaux | France | Centre Hospitalier Universitaire Pellegrin Tripode | | 0556795679 | Neurology |
| NAVARRO | Vincent | Paris | France | Centre Hospitalier Universitaire de la Pitié-Salpêtrière | | 0142161813 | Neurology |
| RHEIMS | Sylvain | Lyon | France | Hospices Civils de Lyon | | 0472357044 | Neurology |
| VALTON | Luc | Toulouse | France | Hôpital Rangueil | | - | Neurology |

### LIST OF COLLABORATORS

#### - PHARMACY

**Dr Christine PIVOT**
**Dr Fabrice PIROT**

Service pharmaceutique
Essais cliniques
Unité de préparation et contrôles des médicaments
Hôpital Edouard Herriot
5, place d'Arsonval
F - 69437 Lyon cedex 03

Tél : + 33 4 72 11 78 74 / Fax: + 33 4 72 11 78 76


#### - IN PARTICIPATING CENTERS

Dr Hélène CATENOIX, Dr Jean ISNARD, Dr Alexandra MONTAVONT, Dr Sébastien BOULOGNE, Service de Neurologie Fonctionnelle et d'Epileptologie, Hospices Civils de Lyon

Dr Agnès TREBUCHON, Dr Ailen McGONIGAL, Dr Sandrine AUBERT, Dr Francesca BONINI, Centre Hospitalier Universitaire de la Timone, Marseille

Dr William SZURHAJ, Dr Arnaud DELVAL, Dr Maxime CHOCHOI, Centre Hospitalier Régional et Universitaire Roger Salengro, Lille

Dr Maria-Paola VALENTI, Hôpitaux Universitaires de Strasbourg, Strasbourg

Dr Jean-Pierre VIGNAL, Pr Louise TYVAERT, Dr Irina KLEMINA, Centre Hospitalier Universitaire de Nancy, Nancy

Dr Marie de Montaudouin, Hôpital Pellegrin Tripode, Bordeaux

Pr Sophie DUPONT, Dr Calude ADAM, Centre Hospitalier Universitaire de la Pitié-Salpêtrière, Paris

Dr Marie DENUELLE, Hôpital Rangueil, Toulouse

#### LIST OF ABREVIATIONS

AED : Antiepileptic Drug
DSMB : Data and Safety Monitoring Board
EMU : Epileptic Monitoring Unit
GTCS : Generalized tonic-clonic seizures
MRI : Magnetic resonance Imaging
PET : Positron Emission Tomography
SPECT : Single-photon emission computed tomography
SUDEP : Sudden unexpected death in epilepsy
WHO : World Health Organization

| **Study Title:**<br>Efficacy of Naloxone in reducing postictal central respiratory dysfunction in patients with epilepsy |
|---|
| **Context:**<br>Sudden unexpected death in epilepsy (SUDEP) primarily affects young adults with drug-resistant epilepsy, with an incidence of about 0.4%/year. The diagnosis of SUDEP requires that anamnestic data and post-mortem examination do not reveal a structural or toxicological cause for death. Generalized tonic-clonic seizures (GTCS) are the main risk factor for SUDEP, which they appear to trigger in most instances. Indeed, experimental and clinical data strongly suggest that most SUDEP result from a postictal respiratory dysfunction progressing to terminal apnea, later followed by cardiac arrest.<br><br>Postictal apnea could partly derive from a seizure-induced massive release of endogenous opioids. Animal studies suggest that such seizure-related release of endogenous opioid peptides participate to termination of seizures. In patients with epilepsy, functional imaging studies have confirmed that seizures induce release of endogenous opioids. The brainstem respiratory centers contain the highest density in opioid receptors, accounting for respiratory depression being one of the cardinal symptoms of opioid overdose.<br><br>Our hypothesis is that SUDEP partly results from a post-ictal apnea promoted by a GTCS-induced massive release of endogenous opioids, and that an opioid antagonist could represent an effective preventive treatment of SUDEP. This could be achieved by chronic administration of Naltrexone, an opioid antagonist that has been used in a large population of patients with chronic alcoholism at high risk of seizures, without showing any pro-convulsant effect. This is a crucial feasibility issue since antagonising a mechanism thought to participate to seizure termination could theoretically aggravate seizures.<br><br>Before evaluating the efficacy of chronic administration of naltrexone, it is legitimate to perform a proof of concept study by testing the acute effect of an equivalent injectable treatment (Naloxone) in the immediate aftermath of GTCS recorded inhospital during video-EEG monitoring of patients with refractory epilepsy. One third of these patients develop postictal respiratory dysfunction and hypoxemia, which should be reduced by our intervention if our hypothesis is correct. |
| **Objectives:**<br>**Principal Objective:**<br>The main objective of the study is to evaluate the efficacy of 0.4 mg intravenous naloxone, versus placebo, administered in the immediate aftermath of a GTCS, in reducing the severity of the postictal central respiratory dysfunction occurring after the end of the seizure, as measured by pulse oximetry.<br><br>**Secondary Objectives:**<br>Assess the impact of naloxone on respiratory parameters such as the frequency of apneas (> 10 seconds) occurring after the end of the seizure. |

Assess the impact of naloxone on 02 administration requirement and on cardiorespiratory rescue procedure requirement after the end of the seizure.

Assess the impact of naloxone on the postictal generalized EEG suppression, the duration of which is correlated with the risk of SUDEP, and the severity of which could also result from a seizure-related release of endogenous opioids.

Assess the impact of naloxone on the duration of the postictal coma following a GTCS, and the time required by the patient to recover preictal clinical condition. In the future, this clinical benefit might be enough significant to justify the systematic use of naloxone after GTCS in inpatients

- Assess the impact of naloxone on the duration of the postictal immobility following a GTCS

Assess the frequency and severity of adverse events related to the treatment with naloxone, such as increased posictal pain and/or early recurrence of GCTS.

**Endpoints:**

**Primary Endpoint:**

Delay between the end of the seizure and recovery of oxygen saturation ($SpO_2$) ≥ 90%

**Secondary Endpoints:**

- Other respiratory parameters

  - Delay between the naloxone or placebo injection and recovery of oxygen saturation ($SpO_2$) ≥ 90%

  - Proportion of patients whose $SpO_2$ is <70%, between 70 and 80%, between 80% and 85% and between 85% and 90% at least 5 seconds 30 seconds, 1 minute, 2 minutes, 3 minutes, 4 minutes and 5 minutes after the end of the GTCS.

  - Desaturation nadir after the end of the GTCS

- Number of patients in whom $0_2$ administration is required within the ten minutes following the end of a GTCS.
- Number of patients in whom cardiorespiratory rescue procedure is required within the ten minutes following the end of a GTCS
- Total duration of the postictal generalized EEG suppression, defined as lack of detectable EEG activity >10 μV in amplitude on all leads.
- Total duration of the postictal coma, defined as the delay between the end of the seizure and the recovery of consciousness assessed by the ability to meet one single verbal command (handshake).
- Report of adverse events observed throughout the study
- Assessment of pain, using a visual analog scale, immediately after the recovery of consciousness following the postictal coma.
- Total duration of the postictal immobility, defined as the delay between the end of the seizure and the first spontaneous movement of the patient, as assessed on the video recording.
- Number of patients who have a second GCTS within 120 minutes after the intravenous injection.

**Trial design: DOUBLE-BLIND RANDOMIZED PLACEBO-CONTROLLED TRIAL**

About 25% of patients with drug-resistant partial epilepsy who undergo long-term video-EEG monitoring develop at least one partial secondary generalized tonic-clonic seizure. However, these patients cannot be individualized *a priori*. Therefore, all adult patients with drug-resistant epilepsy who will undergo long-term video-EEG monitoring in one of the participating centres, will lack all exclusion criteria, and will give their written informed consent to participate to the study if they develop GTCS, will be included in the study. They will all benefit from continuous monitoring of pulse oximetry (together with video, EEG, and respiration recordings), and will be equipped with a peripheral venous catheter throughout the video-EEG. The modalities of the video-EEG monitoring will be consistent with the current practices and similar across the 8 centres (apart from the venous catheter which is not standard practice).

In case of occurrence of a generalized tonic-clonic seizure, patients will be randomized (1:1) to receive intravenous naloxone (0.4 mg) or placebo. Placebo will be isotonic sodium chloride which preparation and packaging will be centralized to ensure its indistinguishability from naloxone. Randomization will be centralized and stratified by centre. The evolution from a partial seizure to a GTCS being gradual, and the total duration of the seizure ranging from 2 to 3 minutes, the injection will be prepared during the course of the seizure. Given the assumptions about the role of endogenous opioids release in the spontaneous termination of seizures, naloxone will be administrated immediately after the end of the GTCS and not before.

All digital data (video, EEG, respiration, $SpO_2$) will be centralized and evaluated blind to other data by the PI of the study who will not be involved in the video-EEG monitoring of the included patients. The same automatic and objective analysis of $SpO_2$ data than the one already developed in the PHRC REPOMSE will be performed.

**Number of patients:**

Sample size was revised using a log rank test with a two-sided alternative hypothesis. For a significance level of 5% (two-tailed), assuming a hazard ratio of 2.414 calculated, i.e based on an effect size similar to the effect size of oxygenotherapy in a delay of 60 seconds (Rheims et al., 2019), 40 events should be observed to reject the null hypothesis in 80% of cases,. A proportion of 89% of patients being expected to recover SpO2 ≥ 90% 120 seconds after the end of the seizure and considering a proportion of unusauble records for technical reasons of 20%, at least 54 patients should be randomized.

Assuming that about 10% of patients with drug-resistant partial epilepsy who undergo long-term video-EEG monitoring develop at least one partial secondary generalized tonic-clonic seizure, a total of 554 patients will be included in the study.

**Recruitment Procedures:**

All adult patients with drug-resistant epilepsy who will undergo long-term video-EEG monitoring in one of the participating centres, will lack all exclusion criteria, and will give their written informed consent to participate to the study, will be included in the study. Among them, those who will develop supervised GTCS during the monitoring will be randomized and will receive the treatment.

If a patient is already included but non-randomized needs a new long-term video-EEG monitoring, the protocol will be proposed again. The investigator will explain the study again and the patient will give his written informed consent again.

**Patients:**

**Inclusion Criteria:**

<u>For inclusion</u>

Adult patient (≥ 18 years) suffering from drug-resistant partial epilepsy

Patient undergoing long-term video-EEG monitoring in one of the participating centre to record and characterize its seizure

Patient who gave its written informed consent to participate to the study, <u>For randomization</u>

Patient who suffers a secondary generalized tonic-clonic seizure during the long-term video-EEG monitoring while being supervised by a nurse or a physician

**Exclusion Criteria:**

Age < 18 years

Patient that has already been randomized in this study

Pregnant or breastfeeding women

| |
|---|
| Hypersensitivity to naloxone<br>History of severe heart disease (myocardial infarction, heart failure disorder, arrhythmia severe hypertension)<br>Ongoing opioïd treatment, including both pure agonists and partial agonists<br>Addiction to opioïds, heroin, or any similar substance<br>Patient participating in another drug trial for less than 2 months |
| **Test Product:**<br>In case of occurrence of a generalised tonic-clonic seizure, patients will be randomized (1:1) to receive intravenous naloxone (0.4 mg), an opioid agonist without partial agonist activity, or placebo |
| **Total Maximum Study Duration**:<br>Inclusion period: 66 months<br>Follow-up period per patient: 36 days |
| **Expected Impact**:<br>SUDEP has become a major issue for patients with epilepsy and their physicians. A recent study (Sillampa and Shinnar NEJM 2010) reported that up to 20% of patients with childhood onset drug resistant epilepsy will die of a SUDEP by the age of 45. No preventive treatment is available and the development of new therapeutic approaches has been prioritised by the WHO. Given the pathophysiological link between the occurrence of central apnea in the aftermath of GTCS and the risk of SUDEP, treatment strategies aiming at reducing the severity of postictal respiratory dysfunction appears as one of the most promising way to prevent SUDEP.<br><br>The demonstration of naloxone efficacy on the severity of postictal hypoxemia will have two primary consequences. First, naloxone would be the first and only therapeutic approach which could be immediately delivered to reverse postictal apnea, especially during long-term video-EEG monitoring. Although rare, SUDEPs have occurred during video-EEG in several epilepsy monitoring units in Europe, including few French centres. In addition, the availability of intramuscular route for naloxone renders possible its use at home, especially for patients in whom severe postictal hypoxemia would have been observed inhospital. By reducing postictal apnea, naloxone might also shorten the postictal phase and promote quicker recovery of consciousness.<br><br>Second, the demonstration that an opioid antagonist can effectively reduce postictal apnea would pave the way for an assessment of a preventive therapy targeting the same pathophysiological pathway using Naltrexone. Indeed, this orally administered opioid antagonist can be delivered long term with an excellent safety profile, including in patients undergoing alcohol withdrawal at high risk of seizure, in whom pro-convulsive effect has not been observed. Following this naloxone study, the impact of chronic naltrexone might first be tested on postictal hypoxemia during video-EEG (by treating patients chronically before and during their video-EEG monitoring). If this second study proved positive, large scale study in ambulatory patients could then be undertaken to test the impact of chronic naltrexone on the risk of SUDEP in high risk population. |
| **Research Sites**:<br>8 French Epilepsy Monitoring Units from the National Research Network on SUDEP predictors (PHRC National REPOMSE 2009)<br>Marseille, Lille, Strasbourg, Nancy, Bordeaux, Paris, Toulouse and Lyon |
| **Sponsor:**<br>Hospices Civils de Lyon, 3 Quai des Célestins, F - 69229 Lyon cedex 02<br>Tel.: +33 4 72 40 68 40 Fax : + 33 4 72 11 51 90<br> |
| **Coordinator investigator:**<br>Service de Neurologie Fonctionnelle et d'Epileptologie et Institut des Epilepsies, Hôpital Neurologique<br>Hospices Civils de Lyon, 59 boulevard Pinel, F - 69677 Bron Cedex<br>Tel.: + 33 4 72 35 70 44 / Fax: + 33 4 72 11 80 39 |

| |
|---|
| |
| **Methodology, coordination and data analysis** **:**<br>Unité de Recherche Clinique (Pôle IMER) – Hospices Civils de Lyon<br>162 avenue Lacassagne, F - 69424 Lyon cedex 03<br>Tel.: +33 4 72 11 51 68<br> |
| Favorable opinion from the ethics committee (CPP SUD EST II), on 22/10/2014 |
| Authorization from the competent authorities, on 03/11/2014 |
| EUDRACT number: 2014-003003-30 |

### 1 CONTEXT AND JUSTIFICATION

Our project aims to develop a new therapeutic approach of seizure-related respiratory dysfunction, a frequent complication of epilepsy which currently benefits from no specific treatment although its occurrence might be associated with risk of sudden unexpected death in epilepsy (SUDEP). In this context, we would like to evaluate the efficacy of an opioid antagonist, naloxone, in reducing post-ictal central apnea in the immediate aftermath of generalized tonic-clonic seizures (GTCS) in patients with drug-resistant partial epilepsy undergoing long term video-EEG recording in Epilepsy Monitoring Units (EMUs). The rationale of this proposal is based on the following arguments, which will be further detailed in the next chapters.

1) SUDEP primarily affects young adults with drug-resistant epilepsy, with an incidence of about 0.4%/year. There is currently no effective treatment to prevent SUDEP, apart from optimising antiepileptic drugs (AEDs). As underscored by the World Health Organization (WHO), there is an urgent need to develop specific therapeutic approaches to tackle this issue.

2) GTCS are the main risk factor for SUDEP, which they appear to trigger in most instances.

3) Experimental and clinical data strongly suggest that most SUDEP result from a postictal respiratory dysfunction progressing to terminal apnea, later followed by cardiac arrest. Furthermore, it has been shown that one third of patients with drug-resistant partial epilepsy develop transient respiratory dysfunction and hypoxemia during seizures. Given the pathophysiological link between the occurrence of postictal central apnea and the risk of SUDEP, treatment strategies aiming at reducing the severity of postictal respiratory dysfunction appears as one promising way to prevent SUDEP.

4) Animal studies suggest that seizure-related release of endogenous opioid peptides participate to termination of seizures. In patients with epilepsy, functional imaging studies have confirmed that seizures induce release of endogenous opioids.

5) Breathing is generated by a complex neuronal network in the brainstem. Opioids inhibit activity of respiratory neurons in the medulla, leading to severe respiratory depression, as observed in opioid overdose.

6) Our hypothesis is that SUDEP partly results from a post-ictal apnea promoted by a GTCS-induced massive release of endogenous opioids, and that an opioid antagonist could represent an effective preventive treatment of SUDEP. This could be achieved by chronic administration of Naltrexone, an opioid antagonist that has been used in a large population of patients with chronic alcoholism at high risk of seizures, without showing any pro-convulsant effect. This is a crucial feasibility issue since antagonising a mechanism thought to participate to seizure termination could theoretically aggravate

seizures. Before evaluating the efficacy of chronic administration of naltrexone, it is legitimate to perform a proof of concept study by testing the acute effect of an equivalent injectable treatment (Naloxone) in the immediate aftermath of GTCS recorded inhospital during video-EEG monitoring of patients with refractory epilepsy.

#### 1.1 Drug-resistant focal Epilepsy: definition, epidemiology and management

According to the International League Against Epilepsy, drug-resistant focal epilepsy (DRFE) is defined as an epileptic disorder in which semiology or findings at investigation disclose a localized origin of the seizures and in which seizures are not controlled after adequate trials of two tolerated, appropriately chosen and used antiepileptic drug schedules (ILAE, 1989; Kwan et al., 2010).

Among the 50 to 60 million individuals suffering from epilepsy worldwide, up to one third might develop drug-resistance (Kwan and Brodie, 2000). In France, this represents about 150,000 patients and 10,000 new cases each year (Jallon, 2004).

In this population, epilepsy surgery represents the optimal treatment option (Schuele and Luders, 2008). Patient selection for epilepsy surgery is a two-step procedure that first aims to identify potential surgical candidates who should benefit from a presurgical evaluation, and then to determine in each assessed individual whether the risk/benefit ratio for surgery is acceptable (Ryvlin and Rheims, 2008). This evaluation primarily requires localizing the epileptogenic zone (EZ), ie the minimum amount of brain tissue that should be resected to render the patient seizure-free. EZ should be unique and not overlapping with eloquent brain regions. Conclusion about the patient's operability and the chance of successful epilepsy surgery results from the integration of a large set of data. This process can be divided into two stages (Ryvlin and Rheims, 2008). After each step, the medical team evaluates whether or not it is legitimate to continue the presurgical evaluation as well as the patient's operability. The first stage - Phase I exploration - is mandatory, and includes neuropsychological assessment, morphological assessment (MRI) and long-term video-EEG monitoring of one to two weeks duration. This latter allows to capture patient's seizures with simultaneous EEG, but also heart rate and respiratory rate. In addition, all EMUs participating to our project systematically couple video-EEG with recording of oxygen saturation with pulse oximetry. In some patients, video-EEG might be supplemented by various techniques of functional imaging (Positron Emission Tomography, SPECT, fMRI). Overall, data obtained during phase I exploration are strong enough to propose surgical procedure in about 50% of patients eligible for epilepsy surgery. In the remaining patients, a second stage - exploration phase II – is required to further clarify the localization of the EZ. This phase II consists in invasive EEG monitoring, using intracerebral electrodes (video-SEEG).

Patients who undergo video-EEG or video-SEEG primarily develop simple or complex partial seizures during the monitoring. However, partial seizures with secondary tonic-clonic generalizations (SGTCS) are not rare. Medical records of the Mayo Clinic Epilepsy Monitoring Unit showed that 24% of patients exhibited at least one SGTCS during long term video-EEG monitoring (Noe and Drazkowski, 2009). Other studies reported similar pictures, with SGTCS corresponding to 26-45% of all recorded seizures (Bateman et al., 2008; Di Gennaro et al., 2012). Few centres however reported lower incidence (Atkinson et al., 2012). In our own experience, about 25% of patients exhibit at least one SGTCS during long-term EEG monitoring.

The study proposed herein will include patients in whom presurgical evaluation is required, and who will undergo long-term video-EEG or video-SEEG monitoring for clinical purpose.

#### 1.2 Sudden unexpected death in epilepsy or SUDEP

##### 1.2.1 Definition

SUDEP is defined as sudden, unexpected, witnessed or unwitnessed, nontraumatic and nondrowning death in patients with epilepsy, with or without evidence for a seizure and excluding documented status epilepticus, in which postmortem examination does not reveal a toxicologic or anatomic cause of death (Nashef et al., 2012).

It should however be noted that ample evidence shows that majority of SUDEP, if not all, occurs in the immediate aftermath of a seizure (Nashef et al., 1998; Langan et al., 2002; Tomson et al., 2008). Thus, a seizure is almost always observed in the few minutes before the SUDEP when this latter occurs in the presence of a witness (Nashef et al., 1998; Langan et al., 2002; Tomson et al., 2008).

##### 1.2.2 Epidemiology and risk factors

Results from a US population-based study indicate that the overall rate of sudden unexpected death in people with epilepsy is more than 20 times higher than in the general population (Ficker et al., 1998). However, the risk of SUDEP varies depending of the type epilepsy, with a significant higher risk in patients with drug-resistant epilepsy (Shorvon and Tomson, 2011). While incidence rates of 0·9 to 2·3 per 1000 person-years have been reported in general epilepsy populations, the incidence of SUDEP is estimated to be 4/1000 patient-years in DRFE (Shorvon and Tomson, 2011), and peaks at 9/1000 patient-years in epilepsy surgery candidates (Dasheiff, 1991), an annual risk equivalent to that of rupture of intracerebral aneurysms (Morita et al., 2012). The incidence of SUDEP in DRFE is the highest in young adults between 18 and 45 years, and is responsible of nearly 500 deaths per year in England, representing a major public health issue (Pedley and Hauser, 2002). Similarly, a recent study reported that up to 20% of patients with childhood onset drug resistant epilepsy will die of a SUDEP by the age of 45 (Sillanpaa and Shinnar, 2010).

Other SUDEP risk factors have been individualized (Shorvon and Tomson, 2011). However, the presence and frequency of generalized tonic–clonic seizures (GTCS; either primary or secondary generalized) was found to represent the main risk factor, with an odds ratio of >15 for patients with three or more GTCS per month (Hesdorffer et al., 2012). A few other risk factors proved significant, but with odds ratios <2, including male gender, age of onset of epilepsy <16 years, duration of epilepsy >15 years, and polytherapy (Hesdorffer et al., 2011). However, when adjustments were made for the number of GTCS, neither polytherapy nor the use of specific antiepileptic drugs (AEDs) such as lamotrigine or carbamazepine, was associated with an increased risk of SUDEP (Hesdorffer et al., 2012). Interestingly, it has also been suggested that nocturnal occurrence of GTCS might be at higher risk of SUDEP, an observation which could either reflect circadian differences in the brain response to GTCS during sleep, with greater risk of neurovegetative dysfunction during the night, or more effective supervision and timely resuscitation during day-time (Ryvlin et al., 2013b).

Overall, it appears from these epidemiological data that the patients who are exposed to the highest risk of SUDEP are young adults who suffer from DRFE with frequent GTCS. Accordingly, it has been suggested that research on SUDEP prevention should focus on this specific population (Hesdorffer et al., 2012; Ryvlin et al., 2013a).

##### 1.2.3 Prevention of SUDEP

There is currently no effective treatment to prevent SUDEP apart from optimising antiepileptic drugs (AEDs) (Ryvlin et al., 2013a). Indeed, the only controlled data available in the field, published by our group, suggest that regular revision of AEDs regimen might be protective (Ryvlin et al., 2011). It has also been suggested that successful epilepsy surgery should offer effective protection against SUDEP. Although this conclusion is supported by studies showing higher risk of SUDEP in patients who failed surgery as compared to those who achieved seizure freedom, we still lack definite proof that this

difference primarily reflects the impact of epilepsy surgery, rather than preexisting biologic differences between the two groups (Ryvlin et al., 2006).
As pointed out by the WHO (WHO, 2010), there is therefore an urgent need to develop specific preventive treatment of SUDEP.

###### 1.2.4 Pathophysiology of SUDEP

The exact pathophysiological mechanisms that lead to SUDEP remain unknown (Devinsky, 2011; Shorvon and Tomson, 2011). However, as detailed below, most of the evidence lends support to the predominant role of central respiratory dysfunction (Tomson et al., 2008; Devinsky, 2011). The possibility that the primary event is an ictal cardiac dysrhythmia or another autonomic or central processess appears less likely, or might account for a minority of SUDEP (Tomson et al., 2008; Devinsky, 2011). For instance, ictal asystole, defined as a sinusal pause of at least 3 seconds occurring during a seizure, usually has a duration of ≤ 60 seconds, and is spontaneously reversible (Scott and Fish, 2000; Rocamora et al., 2003; Rugg-Gunn et al., 2004; Schuele et al., 2007; Tomson et al., 2008). Only rare patients with ictal asystole have undergone cardiopulmonary resuscitation (Lanz et al., 2011). Incidence of ictal asystole in DRFE, estimated at 12 per 100 patient-years, is about 30 times greater than incidence of SUDEP (Scott and Fish, 2000; Rocamora et al., 2003; Rugg-Gunn et al., 2004; Schuele et al., 2007; Tomson et al., 2008).

Thus, both experimental and clinical data strongly suggest that most SUDEP result from a postictal respiratory dysfunction progressing to terminal apnea, later followed by cardiac arrest.

(i) Apnea was the primary cause of death in three animal models of SUDEP, one in sheep, the other two in DBA / 2 and DB1mice suffering from and audiogenic seizures (Johnston et al., 1995; Johnston et al., 1997; Venit et al., 2004; Tupal and Faingold, 2006).
(ii) SUDEP witnesses frequently reported occurrence of respiratory distress in the minutes before death (Nashef et al., 1998; Langan et al., 2002). Moreover, patients who died from SUDEP, typically when they are alone at night in their beds, are frequently found face down in the pillow, suggesting a post-ictal suffocation (Kloster and Engelskjon, 1999).
(iii) We recently conducted a worldwide study funded by the international league against epilepsy which aimed at retrieving data from all monitored cardiorespiratory arrests which had occurred during long-term video-EEG monitoring (Ryvlin et al., 2013b). Detailed analysis of video-EEG and EKG material at the time of cardiorespiratory arrests showed postictal central apnea in all SUDEP. This central respiratory dysfunction typically occurred between 1 and 3

minutes after the end of a GTCS, in combination with severe bradycardia or transient asystole (Ryvlin et al., 2013b). The cardiorespiratory collapse was terminal in one third of patients. In the remainder, it was followed by transient restoration of cardiac function associated with abnormal and possibly ineffective respiration likely aggravated by the prone position. Respiration then progressively deteriorated until terminal apnea which always preceded terminal asystole (Ryvlin et al., 2013b). Most importantly, the short delay between the end of seizure and onset of apnea supports the view that the central respiratory dysfunction is at least partly triggered by the release of neurotransmitter involved in seizure termination (including endogenous opoids), and that there a short window during which this mechanism might be reversed by an appropriate treatment.

Another factor which is associated with SUDEP is post-ictal generalized EEG suppression (PGES), defined as postictal (within 30 seconds), generalized absence of electroencephalographic activity >10mV (Lhatoo et al., 2010). However, the exact pathophysiological significance of PGES remains controversial. The duration of PGES predicted the risk of SUDEP in one study (Lhatoo et al., 2010) but not in another (Surges et al., 2011). Similarly, association between PGES and periictal autonomic changes proved controversial (Poh et al., 2012; Lamberts et al., 2013). PGES was also reported to be associated with more nursing interventions and ictal respiratory dysfunction (Semmelroch et al., 2012; Seyal et al., 2012), but not with postictal apnea (Seyal et al., 2012). According to current knowledge, PGES might primarily represent an ancillary marker of profound postictal cerebral dysfunction, possibly triggered by the same neurobiological mechanisms leading to post-ictal respiratory dysfunction.

Two studies investigated the frequency and nature of per-and post-ictal apneas in adult patients with DRFE undergoing video-EEG monitoring (Nashef et al., 1996; Bateman et al., 2008). Their objective was to study all apneas occurring during seizures, and not only those responsible for acute respiratory distress (Nashef et al., 1996; Bateman et al., 2008). Importantly, these studies have used recording of respiratory parameters, including the measurement of pulse oximetry, showing that this latter remained reliable in the vast majority of seizures, whereas other respiratory parameters, such as those collected from nasal probe, often suffered from artifacts prohibiting their interpretation. Overall, both studies demonstrated two major findings: (i) Two thirds of patients will develop oxygen desaturation <90% during at least one of their recorded seizure (Nashef et al., 1996; Bateman et al., 2008). Apneas occur in 33% of seizures, both in strictly partial seizures and in secondarily generalized tonic-clonic seizures, and can result in oxygen desaturation up to 50% (Bateman et al., 2008). (ii) Oxygen desaturation begins during the seizure, but will often continue or get worse during the post-ictal period (Nashef et al., 1996; Bateman et al., 2008), an evolutive pattern similar to that of central apnea observed in monitored

SUDEP (Ryvlin et al., 2013b). In addition, analysis of concomitant end-tidal $CO_2$ showed elevation parallel to hypoxemia, as observed in alveolar hypoventilation (Seyal et al., 2010). A relation between the occurrence and/or severity of seizure-related apnea and SUDEP has been suggested, but not been formally established (Schuele et al., 2011). This important issue is the primary objective of an ongoing multicenter propective study, based on the French National Research Network on SUDEP predictors coordinated by our team (PHRC National REPOMSE 2009). Nevertheless, available data strongly suggest that focusing on seizure-related central apnea in patients who demonstrate the highest risk of SUDEP, i.e patients with DRFE and GTCS who undergo presurgical evaluation, is a relevant strategy to further investigate the underlying mechanisms of SUDEP and develop new therapeutic approachs (Ryvlin et al., 2013a).

Given the role of serotonin in the regulation of breathing (Richter et al., 2003), serotonergic agents have thus been proposed to reduce postictal apnea with the hope to prevent SUDEP (Ryvlin et al., 2013a). Two randomised controlled studies are currently testing this hypothesis in the USA and in France, the later being coordinated by our centre. Although other key players of the respiratory network regulation might represent interesting target for SUDEP prevention, including the opioid system, no other strategy has been evaluated so far.

#### 1.3 Endogenous opioids and epilepsy

##### 1.3.1 Endogenous opioid system

The endogenous opioid system consists of three families of peptides, b-endorphin, enkephalin and dynorphin, and three families of receptors, $\mu$ (MOR), $\delta$ (DOR), and $\kappa$ (KOP) which are defined pharmacologically by their blockade by naloxone. Two additional endogenous peptides, endormorphin-1 and -2 have been identified, but their function remains incompletely understood (Fichna et al., 2007). Opioid peptides and their receptors have a widespread but selective distribution in the central and peripheral nervous systems (Le Merrer et al., 2009; Benarroch, 2012). The opioid system plays an important role in pain control, drug addiction, modulation of emotion and stress response (Benarroch, 2012). It has also been suggested that it may participate in the pathophysiology of seizures (Benarroch, 2012).

Endogeous opioids result from cleavage of large protein precursors, proopiomelanocortin for $\beta$-endorphin, preproenkephalin for enkephalins and preprodynorphyn for dynorphins (Le Merrer et al., 2009; Benarroch, 2012). Neurons synthesizing $\beta$-endorphin are restricted to two locations, the arcuate nucleus of the hypothalamus and the nucleus tractus solitarius in the medulla (Fichna et al., 2007; Benarroch, 2012). However, they provide widespread projections throughout the central nervous system, including all limbic forebrain and midbrain areas, the brainstem and the spinal cord (Fichna et al., 2007; Benarroch, 2012). In contrast, preproenkephalin and preprodynorphin are expressed in local neurons distributed at multiples levels of the central nervous system, including the neocortex, hippocampus, thalamus, basal ganglia, hypothalamus and medulla (Fichna et al., 2007; Benarroch, 2012).

Endogenous opioids demonstrate different affinities with the three different receptors subtypes, MOR, DOR, and KOR (Fichna et al., 2007; Benarroch, 2012). β-endorphin primarily acts via MOR and DOR, enkephalins via DOR and dynorphins via KOR. MOR is the most abundant receptor in the amygdala, thalamus and the brainstem whereas DOR is mostly expressed in the cerebral cortex, includind limbic areas, and striatum and the KOR in the claustrum, striatum and hypothalamus.

However, activation of all three opioids receptors results in the same physiological effects. Thus, all opioids elicit presynaptic and post-synaptic inhibition through activation of G-protein-coupled receptors which lead to inhibition of adenylyl cyclase, inhibition of voltage-gated calcium channels, particularly presynaptic P/Q- and N-type channels, and activation of inwardly rectifying potassium channels (Trescot et al., 2008). Overall, activation of opioid receptors by their ligands result in presynaptic inhibition of neurotransmitter release, postsynaptic hyperpolarization and decrease in neuronal activity (Trescot et al., 2008).

##### 1.3.2 Endogenous opioids during seizures

Activation of endogenous opioid system during seizures has been suggested by animal studies as well by functional imaging and pathological studies in patients with epilepsy (Henriksen and Willoch, 2008; Benarroch, 2012). Most convincing evidences imply a role for opioids in post-ictal seizure inhibition.

Potentiation of endogenous anti-ictal mechanisms by opioids has been shown in animal models of epilepsy (Tortella et al., 1985). Kainic acid-induced seizures thus elicit dynorphin release in rodent hippocampus and activation of presynaptic KOR inhibits gluatmate release and limits the spread of excitability in this region (Solbrig et al., 2006; Loacker et al., 2007). Similarly, several reports indicate that enkephalin release is induced by epileptiform activity (Rocha et al., 1994) and that MOR activation can induce anticonvulsant effects (Albertson et al., 1984; Bohme et al., 1987). It should however be noted that dual effect of MOR has been suggested with facilitation of the epileptogenesis process but increase of refractoriness of subsequent seizures during the postictal period (Rocha et al., 1991; Rocha et al., 1993; Rocha et al., 1996).

In patients with epilepsy, seizure-related activation of the endogenous opioid system relies on several observations:

(i) Selective changes in MOR binding and functional coupling to G-proteins have been observed in surgical specimens obtained from temporal neocortex of epileptic patients. Although MOR binding was enhanced, activation of G-protein mediated by MOR, and therefore physiological effect of MOR activation, was decreased (Rocha et al., 2009). Similar increased MOR binding has been described in the hippocampus of patients with pharmacoresistant mesial temporal lobe epilepsy (Cuellar-Herrera et al., 2012).

(ii) A functional polymorphism in the prodynorphin gene promotor, which limits gene expression over basal condition, is associated with temporal lobe epilepsy (Stogmann et al., 2002).

(iii) Imaging of opioid receptors with positron emission tomography showed specific alterations both during the interictal period and following epileptic seizures. Interictal PET studies in patients with temporal lobe epilepsy have thus shown increased binding in the lateral temporal neocortex on the side of the epileptogenic focus with the MOP-receptor selective radioligand [11C]carfentanyl (Frost et al., 1988; Mayberg et al., 1991) as well as with the DOP-receptor selective radioligand [11C]methylnaltrindole (Madar et al., 1997) but no side-to-side differences of [11C]diprenorphine binding (Mayberg et al., 1991; Bartenstein et al., 1994). Ictal and immediate post-ictal modifications of opioid receptor binding have been investigated with [11C]diprenorphine in patients with reading-induced epilepsy (Koepp et al., 1998), in absence epilepsy (Bartenstein et al., 1993) as well as in patients with temporal lobe epilepsy (Hammers et al., 2007; McGinnity et al., 2013). Importantly, it should be noted that [11C]diprenorphine binds to all three opioid receptor subtypes (Lee et al., 1999). Scans acquired during reading-induced seizures or absences showed decrease in [11C]diprenorphine binding (Bartenstein et al., 1993; Koepp et al., 1998) whereas that performed 8 hours after temporal lobe seizure demonstrated increase in [11C]diprenorphine binding in ipsilateral fusiform gyri, lateral temporal pole and parahippocampal gyrus (Hammers et al., 2007; McGinnity et al., 2013). Taken together, these results have been interpreted as suggesting that synaptic opioid levels increase at the time of seizures, leading to a reduction in [11C]diprenorphine binding, and that this is followed by a gradual recovery of available surface receptors with an overshoot over basal levels which is detected by PET about 8 h after seizures, with a gradual return to normal or low-normal levels during the interictal phase (Hammers et al., 2007).

Overall, there are convincing data to conclude that endogenous opioids are released during focal and generalized seizures. Though potential correlation between the level of activation of the opioid system and the type of epileptic discharge has not specifically been reported yet, GTCS might be associated with massive release of endogenous opioids. Although this phenomenon might participate to seizure termination and/or prevention of early recurrence, it is unlikely that the overall effect of opioid system activation remains so specific. In contrast, additional neuronal effect of opioids might be anticipated, including analgesic effect and/or impact on respiratory rhythms.

##### 1.3.3 Central control of breathing and impact of opioids

The impact of opioids on the control of respiration is well known (Pattinson, 2008) and respiratory depression is one of the cardinal symptoms of opioid overdose (Boyer, 2012). This effect is related to direct inhibition of respiratory neurons by activation of opioid receptors (Pattinson, 2008; Montandon et al., 2011).

The fundamental drive to respiration is generated in the brainstem and is modulated by various inputs originating from the cortex, subcortical nuclei, the brainstem or peripheral chemoreceptors (Feldman et al., 2003). Respiratory neurons are concentrated in three main brainstem areas: the dorsal respiratory group within the nucleus of the solitary tract, the ventrolateral medulla from the level of the spinal–medullary junction through the level of the facial nucleus (i.e. the ventral respiratory column), and in the pontine respiratory group within the dor- solateral pons. These aggregates of brainstem respiratory neurons are interconnected, and together with respiratory-related sensory afferents, are collectively responsible for the automatic control of breathing as well as adaptive changes in breathing to homeostatic and environmental challenges (Feldman et al., 2003).

Within this complex network, a region of the ventro-lateral medulla, the preBötzinger complex (PreBötC), plays an essential role in generating the basic respiratory rhythm. The PreBötC has been individualized both in rodents (Smith et al., 1991; Gray et al., 1999) and in Humans (Lavezzi and Matturri, 2008). It contains excitatory neurokinin-1 receptor (NKR1)-expressing neurons which fire spontaneously and simultaneously with a preinspiratory firing pattern (Feldman et al., 2003). PreBötC neurons send projections to most respiratory-related nuclei in the brainstem (Tan et al., 2010) and their activity controls generation of rhythmic respiratory movements both in vitro and in vivo (Smith et al., 1991; Gray et al., 1999; Tan et al., 2008). Accordingly, specific destruction or silencing of NKR1 neurons induces ataxic breathing or outright central apneas in freely behaving adult rats (McKay et al., 2005;

Tan et al., 2008; Montandon et al., 2011). Specifically, slow toxin-induced neurodegeneration of NKR1 neurons results in an altered respiratory rhythm characterized by ataxic rhythm during wakefulness and apnea during sleep (Gray et al., 1999; McKay et al., 2005), while rapid silencing of these neurons induces a persistent apnea without any respiratory movements to rescue breathing (Tan et al., 2008). Therefore, though slow alteration of PreBötC could partly be compensated by neuronal reorganization, sudden depression of NKR1 neurons activity can lead to serious, even fatal, respiratory failure. Opioid receptor, specifically MOR, are densely expressed in the brainstem respiratory centers (Pattinson, 2008). In neonatal rodents, NKR1 and MOR are coexpressed in PreBötC neurons and application of opioids to the PreBötC slows respiratory rate in vitro (Gray et al., 1999). Furthermore, local application of opioids at the PreBötC in the adult rat in vivo causes respiratory depression or fatal apnea (Montandon et al., 2011).

Alterations of PreBötC neurons, which might lead to breathing disorders or sudden death, have also been observed in pathological specimens (McKay et al., 2005; Lavezzi and Matturri, 2008). Pathological studies of NKR1 neurons in fetuses, newborns and infants showed structural and/or functional alterations in 47% of sudden deaths, and abnormalities of PreBötC migh be involved in the pathophysiology of sudden infant death syndrom (Lavezzi and Matturri, 2008). It has also been suggested that sleep-disordered breathing observed in individuals with Parkinson disease or multiple system atrophy might be related to depletion in NK1R neurons (McKay et al., 2005).

No pathological study of brainstem respiratory networks, and specifically of PreBötC and/or alteration of opioid signaling, has been conducted in SUDEP yet. However, some indirect arguments suggest that opioid-induced breathing impairment could be involved in post-GTCS respiratory dysfunction and SUDEP:

(i) As previously discussed, detailed analysis of respiratory patterns in monitored SUDEP demonstrated a progressive deterioration of respiratory rate until terminal apnea (Ryvlin et al., 2013b). Similarly, respiratory pattern in patients exposed to high doses of opioids demonstrate alteration of rhythmic breathing, with irregular respiratory pattern and/or transient apnea, rather than change in tidal volume (Pattinson, 2008).

(ii) Arousal level, which is profoundly altered during the immediate aftermath of GTCS, critically impacts generation of respiratory rhythms. Wakefulness thus provides a significant drive to the respiratory system. As a matter of fact, deterioration of breathing related to slow destruction of NKR1-expressing PreBötC neurons first appears in sleep and is eventually more pronounced during sleep than wakefulness (McKay et al., 2005). Beyond the voluntary control of ventilation which necessarily involves cortical and subcortical motor networks (McKay et al., 2008), vegetative regulation of respiratory rhythms requires widely distributed systems which are not solely localized within the brainstem. Functional MRI studies in Humans have shown that chemo-stimulated increases in ventilation is not only mediated by brainstem respiratory centers, but involves a large network which includes basal ganglia, thalamus, red nucleus, cerebellum, parietal and cingulate cortices (McKay et al., 2010). Accordingly, the magnitude of opioid-induced respiratory rate suppression is significantly correlated with the degree of cortical activation, with this suppression being more pronounced during deep non-REM sleep or anesthesia (Montandon et al., 2011).

Overall, considering that seizures induce release of endogenous opioids on the one hand and the depressive effect of opioid receptors activation on respiratory neurons on the other, we propose that SUDEP might partly result from a post-ictal apnea promoted by a GTCS-induced massive release of endogenous opioids.

##### 1.4 Acute reversion of potential postictal opioids-induced respiratory depression

Clinical effects of opioids can be reversed and/or prevented by application of antagonists of opioid receptors. Based on the above rational, we therefore propose that an opioid antagonist could represent an effective preventive treatment of SUDEP, a hypothesis that has never been investigated yet.

Two main compounds are available in clinical practice, naloxone and naltrexone.

Naloxone is the gold-standard competitive antagonist of opioid receptors. Blockade by naloxone pharmacologically defines the opioid receptors family. It is active when the parenteral, intranasal, or pulmonary route of administration is used but has negligible bioavailability after oral administration because of extensive first-pass hepatic metabolism (Boyer, 2012). Intravenous administration is approved in France since 1977 for the treatment of acute opioid overdose. In this indication, the recommended starting dose is 0.4 mg, though the effective dose depends on the amount of opioid analgesic the patient has taken or received (Boyer, 2012). In acute opiate overdose, the delay between intravenous injection and awakening ranges from 30 seconds to 2 minutes and its apparent duration of action is 20 to 90 minutes (Boyer, 2012). There is no major safety issue reported with naloxone. Risk of pulmonary edema has been discussed although this latter is present in nearly all fatal cases of opioid overdose, including those that occurred before the development of naloxone (Boyer, 2012). The other side effects observed in context of use of naloxone in patients with opioid dependence are signs of opioid abstinence (e.g., yawning, lacrimation, piloerection, diaphoresis, myalgias, vomiting, and diarrhea), which are unpleasant but not life-threatening (Boyer, 2012).

Naltrexone is a competitive antagonist of MOR and KOR which is primarily used in the chronic management of alcohol dependence and opioid dependence (Ray et al., 2010). It was approved for the treatment of alcohol dependance in 1994. It is usually given by oral route at a standard dose of 50 to 100 mg/day (Ray et al., 2010), though intra-muscular extended-release formulation has been evaluated (Gastfriend, 2011). As with naloxone, no life-threatening side effect has been reported with naltrexone (Ray et al., 2010). The main reason for discontinuation in opioid-dependant patients is nausea, observed in up to 15% of patients (Ray et al., 2010). Although historic investigations with high doses of naltrexone such as 300 mg per day reported a significant increase in hepatic enzymes, systematic reviews concluded that in patients treated for heroin or alcohol addiction, there is no evidence that naltrexone causes clinically significant liver disease or exacerbates serious pre-existing liver disease (Ray et al., 2010).

Neither naloxone nor naltrexone has been specifically investigated in patients with epilepsy (Ray et al., 2010; Boyer, 2012). Antagonising a mechanism thought to participate to seizure termination could theoretically aggravate seizures. It should however be noted that no epilepsy-related alert has been reported for these drugs, even in patients with chronic alcoholism who suffer high risk of seizures (Samokhvalov et al., 2010). Nevertheless, before evaluating the efficacy and safety of chronic administration of naltrexone, it is legitimate to perform a proof of concept study by testing the acute effect of an equivalent injectable treatment (Naloxone) in the immediate aftermath of GTCS recorded inhospital during video-EEG monitoring of patients with refractory epilepsy.

### 2 OBJECTIVES OF THE STUDY

#### 2.1 PRINCIPAL

The main objective of the study is to evaluate the efficacy of 0.4 mg intravenous naloxone, versus placebo, administered in the immediate aftermath of a GTCS, in reducing the severity of the postictal central respiratory dysfunction occurring after the end of the seizure, as measured by pulse oximetry.

#### 2.2 SECONDARY

- Assess the impact of naloxone on respiratory parameters such as the frequency of apneas (> 10 seconds) occurring after the end of the seizure.

- Assess the impact of naloxone on 02 administration requirement and on cardiorespiratory rescue procedure requirement after the end of the seizure.
- Assess the impact of naloxone on the postictal generalized EEG suppression, the duration of which is correlated with the risk of SUDEP, and the severity of which could also result from a seizure-related release of endogenous opioids.
- Assess the impact of naloxone on the duration of the postictal coma following a GTCS, and the time required by the patient to recover preictal clinical condition. In the future, this clinical benefit might be enough significant to justify the systematic use of naloxone after GTCS in inpatients.
- Assess the impact of naloxone on the duration of the postictal immobility following a GTCS
- Assess the frequency and severity of adverse events related to the treatment with naloxone, such as increased posictal pain and/or early recurrence of GCTS.

### 3 ENDPOINTS

#### 3.1 PRIMARY ENDPOINT

Delay between the end of the seizure and recovery of oxygen saturation ($SpO_2$) ≥ 90%

*The primary endpoint, as well as all secondary endpoints, will be assessed in patients who develop a partial secondary generalized seizure during long-term video-EEG/video-SEEG performed for clinical purpose in the presurgical evaluation. In patients who would develop several SGTCS during the monitoring, only one SGTCS will result in randomization, treatment allocation and assessment of outcomes. The modalities of the video-EEG/video-SEEG monitoring will be consistent with the current practices and not modified by the protocol, apart from the venous catheter, required to administer the intervention, which is not standard practice. Systematic recording of pulse oximetry coupled with video-EEG/video-SEEG is standard pratice in all partipating EMUs.*

*The evolution from a partial seizure to a GTCS being gradual, and the total duration of the seizure ranging from 2 to 3 minutes, the injection will be prepared during the course of the seizure and administered immediately after the end of the seizure. As detailed in section 7.3, the procedure for drug preparation and administration, including the randomization process, will be organized to allow intravenous injection within the two minutes following the end of the seizure.*

*The delay between the end of the seizure and administration of the treatment will be precisely determined by the video recording of the event, given that the nurse or physician who will administer the study drug will be required to both loudly tell "début d'injection" and raise an arm at onset of injection.*

*The studies which investigated the frequency and nature of per-and post-ictal apneas in patients with DRFE undergoing video-EEG monitoring showed that oxygen desaturation began during the seizure, but often continued or get worse during the post-ictal period* (Nashef et al., 1996; Bateman et al., 2008)*. In addition, detailed analysis of video-EEG and EKG material in monitored cardiorespiratory arrests showed that central respiratory dysfunction typically occurred between 1 and 3 minutes after the end of a GTCS, in combination with severe bradycardia or transient asystole and PGES* (Ryvlin et al., 2013b)*. The cardiorespiratory collapse was terminal in one third of patients. In the remainder, it was followed by transient restoration of cardiac function associated with abnormal and possibly ineffective respiration likely aggravated by the prone position. Respiration then progressively deteriorated until terminal apnea which always preceded terminal asystole* (Ryvlin et al., 2013b).

*The same automatic and objective analysis of $SpO_2$ data than the one already developed in the PHRC REPOMSE will be performed (see section 8.3).*

*As stated in section 11, the primary endpoint, as well as all secondary endpoints, will primarily be analysed in an intention to treat manner in all randomized patients. However, the analysis will also be reprocessed in the "per-protocol population" of which will be excluded randomized patients having major protocol deviation such as patients in whom the study drug has been administered > 120 seconds after the end of the seizure.*

#### 3.2 SECONDARY ENDPOINTS

- Other respiratory parameters- Delay between the naloxone or placebo injection and recovery of oxygen saturation ($SpO_2$) ≥ 90%

- Proportion of patients whose $SpO_2$ is <70%, between 70 and 80%, between 80% and 85% and between 85% and 90% at least 5 seconds, 30 seconds, 1 minute, 2 minutes, 3 minutes, 4 minutes and 5 minutes after the end of the GTCS.

*Previous studies have shown that the measurement of pulse oximetry is reliable in the vast majority of seizures, including GTCS* (Nashef et al., 1996; Bateman et al., 2008)*. The criterion of hypoxemia duration of at least 5 seconds is however required to eliminate artifactual changes of $SpO_2$.*

- Desaturation nadir after the end of the GTCS

- Number of patients in whom $0_2$ administration is required within the ten minutes following the end of a GTCS.

*Given the potential aggravating effect of prolonged hypoxemia on the mechanisms which lead to central respiratory dysfunction* (Ryvlin et al., 2013b)*, all patients who show Sp02 < 85% during > 2 minutes following the injection of the study drug will receive $0_2$ using high concentration breathing mask (15 L/min).*

- Number of patients in whom cardiorespiratory rescue procedure is required within the ten minutes following the end of a GTCS

*In patients who will demonstrate persistent respiratory distress and oxgen desaturation despite $0_2$ inhalation using high concentration breathing mask (15 L/min), cardiorespiratory rescue procedure will be initiated to immediately manage the risk of cardiorespiratory arrest (see section 8.3)*

- Total duration of the postictal generalized EEG suppression, defined as lack of detectable EEG activity >10 μV in amplitude on all leads.

*This ouctome will only be assessed in patients undergoing scalp Video-EEG. Given the limited brain sampling of invasive recordings, we cannot discrimate postictal focal EEG suppression from generalized EEG suppression in patients undergoing video-SEEG. In addition, the cutoff value which would define absence of electroencephalographic activity with intracranial electrodes remains unknown.*

- Total duration of the postictal coma, defined as the delay between the end of the seizure and the recovery of consciousness assessed by the ability to meet one single verbal command (handshake for exemple).

*This outcome will be assessed using standardised observation and report from the nurse or physician who performed the postictal intravenous injection, confirmed with video recording of the event.*

- Total duration of the postictal immobility, defined as the delay between the end of the seizure and the first spontaneous movement of the patient, as assessed on the video recording.

- Report of adverse events observed throughout the study

- Assessment of pain, using a visual analog scale, immediately after the recovery of consciousness following the postictal coma.

- Number of patients who have a second GCTS within 120 minutes after the intravenous injection.

- Other parameters assessed during the study

  - Age
  - Epilepsy duration
  - Total number of seizures (ie partial seizures and GTCS) over the past three months
  - Number of GTCS over the past three months
  - Localization of the seizure onset zone, as assessed by video-EEG and/or video-SEEG data
  - Body mass index

### 4 CHARACTERISTICS OF THE STUDY

#### 4.1 TRIAL DESIGN

This is a prospective, double-blind, randomised multicentric trial.

About 25% of patients with drug-resistant partial epilepsy who undergo long-term video-EEG/video-SEEG monitoring develop at least one partial secondary generalized tonic-clonic seizure. However, these patients cannot be individualized *a priori*. Therefore, all adult patients with drug-resistant epilepsy who will undergo long-term video-EEG/video-SEEG monitoring in one of the participating centres, will lack all exclusion criteria, and will give their written informed consent, to participate to the study if they develop GTCS, will be included in the study. They will all benefit from continuous monitoring of pulse oximetry, and will be equipped with a peripheral venous catheter throughout the video-EEG/video-SEEG.

In case of occurrence of a generalised tonic-clonic seizure, patients will be randomized (1:1) into two groups:

- Experimental group: patients with drug-resistant partial seizures who suffer a secondary generalised tonic-clonic seizure during long-term video-EEG monitoring and in whom naloxone is administered intravenously immediately after the end of the seizure.
- Control group: patients with drug-resistant partial seizures who suffer a secondary generalised tonic-clonic seizure during long-term video-EEG monitoring and in whom placebo is administered intravenously immediately after the end of the seizure.

#### 4.2 DURATION OF THE STUDY

**Duration of participation of each patient:**
Duration of participation of each patient in the trial will be of maximum 36 days*.

**Anticipated Duration of Recruitment**
66 months

**Long-term video-EEG/video-SEEG monitoring typically lasts between 7 and 15 days, and occasionnaly up to 21 days. However, the total duration of monitoring will be variable across patients, this latter being defined by clinical parameters, specifically the occurrence of a suffisant number of seizures to allow conclusion about the localization of the seizure onset zone. A phone visit will be performed 15 days after the end of the monitoring for safety reasons (see section 8.3).*

### 5 EXPECTED PATIENT OR PUBLIC HEALTH BENEFIT

Although SUDEP is the leading cause of death in people with chronic refractory epilepsy, there is currently no effective treatment to prevent SUDEP, apart from optimising AEDs (Devinsky, 2011; Ryvlin et al., 2011). As underscored by the World Health Organization, there is an urgent need to develop specific therapeutic approaches to tackle this issue. Given the pathophysiological link between the occurrence of central apnea in the aftermath of GTCS and the risk of SUDEP, treatment strategies aiming at reducing the severity of postictal respiratory dysfunction appears as one of the most promising way to prevent SUDEP (Ryvlin et al., 2013a).

The demonstration of naloxone efficacy on the severity of postictal hypoxemia will have two primary consequences:

- First, naloxone would be the first and only therapeutic approach which could be immediately delivered to reverse postictal apnea, especially during long-term video-EEG monitoring. Although rare, SUDEPs have occurred during video-EEG in several epilepsy monitoring units in Europe, including few French centres (Ryvlin et al., 2013b). In addition, the availability of intramuscular route for naloxone (Boyer, 2012) renders possible its use at home, especially for patients in whom severe postictal hypoxemia would have been observed inhospital. By reducing postictal apnea, naloxone might also shorten the postictal phase and promote quicker recovery of consciousness.

- Second, the demonstration that an opioid antagonist can effectively reduce postictal apnea would pave the way for an assessment of a preventive therapy targeting the same pathophysiological pathway using Naltrexone. Indeed, this orally administered opioid antagonist can be delivered long term with an excellent safety profile, including in patients undergoing alcohol withdrawal at high risk of seizure, in whom pro-convulsive effect has not been observed (Ray et al., 2010; Samokhvalov et al., 2010). Following this naloxone study, the impact of chronic naltrexone might first be tested on postictal hypoxemia during video-EEG (by treating patients chronically before and during their video-EEG monitoring). If this second study proved positive, large-scale study in ambulatory patients could then be undertaken to test the impact of chronic naltrexone on the risk of SUDEP in high-risk population.

### 6 SUBJECT SELECTION AND WITHDRAWAL

#### 6.1 RECRUITMENT AND FEASIBILITY

According to the fact that the patients who demonstrate at least one partial seizure with secondary tonic-clonic generalization during long-term video-EEG monitoring cannot be individualized a priori, the feasibility of the study is based on two parameters: (i) the ability to include the eligible population which will consist of patients with drug-resistant partial epilepsy who undergo long-term video-EEG/video-SEEG monitoring and who might demonstrate at least one GTCS during the monitoring, (ii) the proportion of patients from the included population who will suffer at least one GTCS during the monitoring while being supervised and will be effectively randomized.

8 centres will participate to the recruitment (Marseille, Rennes, Saint-Etienne, Montpellier, Tours, Lille, Strasbourg, Grenoble, Nancy, Bordeaux, Paris, Tain l'Hermitage, Lyon, Dijon and Toulouse). All of them are included in the French National Research Network on SUDEP predictors and participate to the REPOMSE study, which primary objective is to study the relation between the per/postictal apnea and the risk of SUDEP and which was granted by the PHRC national in 2009. The modalities of the video-EEG monitoring and patients' management are similar in all participating centers, including modalities

of invasive recording using SEEG in the nine EMUs which perform both video-EEG and video-SEEG monitorings (Marseille, Rennes, Lille, Strasbourg, Grenoble, Nancy, Paris, Lyon and Toulouse). Specifically, all of them benefit from systematic recording of pulse oximetry coupled with video-EEG.

The feasibility of the recruitment of the included population for each centre has taken into account the proportion of patients who fulfil the inclusion and exclusion criteria among those admitted in epilepsy monitoring units and the annual leave (most EMUs close a few weeks every year).

The 15 participating centers totalise 30 beds devoted to long-term video-EEG monitoring for presurgical assessment of drug-resistant partial epilepsy. The average length of stay in these beds is about 10 days, leading to potentially 30 patients per bed per year (900 patients for 30 beds in 15 centers). However, some of these beds are also used for other purpose, such as sleep diseases, and certain eligible patients in our protocol are likely to refuse to participate. Accordingly, the REPOMSE study included 900 patients over a period of 30 months, using inclusion criteria similar to that proposed in this project. Given the effective recruitment of the centres involved in the PHRC REPOMSE and the proportion of patients who usually refuse to participate to randomized trial, we think that inclusion of 700 patients is feasible.

Some studies evalatued the frequency of SGTCS during video-EEG monitoring. Medical records of the Mayo Clinic Epilepsy Monitoring Unit showed that 24% of patients exhibited at least one SGTCS during long term video-EEG monitoring (Noe and Drazkowski, 2009). Other studies reported similar pictures, with SGTCS corresponding to 26-45% of all recorded seizures (Bateman et al., 2008; Di Gennaro et al., 2012). Few centres however reported lower incidence (Atkinson et al., 2012). A preliminary analysis of the data obtained in the REPOMSE study shows that the frequency of GTCS is about 25% of patients. Overall, we estimated that 25% of the included population wil be eligible for randomization (i.e. 0,25 * 700 = 175).

Many patients come back a few months after their first monitoring or for a second monitoring or for a SEEG because they had no seizure during the first one,.

If the patient has been already included but not randomized, patient who come back for a long-term video-EEG monitoring or SEEG monitoring, the protocol will be proposed again. The investigator will explainthe study again and the patient will give his written informed consent again. This procedure will not impact the statistics concerning efficacy and safety, since randomized patient can not be included twice.

ENALEPSIE study might be conducted in parallel with another study in 11 of the 15 participating centers (Bordeaux, Grenoble, Lille, Lyon, Marseille, Montpellier, Nancy, Paris, Rennes, Tain l'Hermitage, Toulouse, Dijon). This other study, entitled Safety of Antiepileptic drug withdrawal in long-term Video-EEG monitoring (SAVE), has been submitted for grant application and is currently under review. SAVE study aims at assessing the impact of a standardized protocol of AEDs withdrawal during long-term VEEG monitoring on the frequency of seizure-related serious adverse events (SAEs) occurring during these monitorings, using a randomized, controlled, parallel group, open-label design, with a cluster randomization, ie applied at the level of centers rather than at the level of patients. Half of participating EMUs will thus be randomized to the group where the standardized protocol of AEDs withdrawal will be used systematically, while the other EMUs will continue their current non-standardized practice of AEDs withdrawal, and will serve as a control group. If SAVE study is granted, it will be implemented in the partipating centers over the same period as ENALEPSIE study. However, we consider that SAVE study should not significantly impact the feasibility of ENALEPSY study for the following reasons:

- ENALEPSIE and SAVE studies differ in tested interventions and evaluated outcomes. Specifically, SAVE study will only test the impact of change in AEDs withdrawal on the occurrence of seizure-related SAEs during videoEEG/videoSEEG. However, it will not evaluate any active intervention in case of occurrence of serious adverse event. As a matter of fact, occurrence of persistant respiratory distress and/or cardiorespiratory arrest in patients included in SAVE study will be managed using the same cardiorespiratory rescue procedure as that planned in ENALEPSIE (see section 8.3). In this context, SAVE study will not include any intervention or procedure which might interfer with the administration of naloxone, its pharmacological effect or the evaluation of its efficacy/safety through the outcomes detailed in the section 3 of the present protocol. In other words, SAVE will interfere with patients' management prior to the occurrence of SGTCS, while ENALEPSIE will interfere with patients' management immediately following a seizure.

- While SAVE study will not affect the evaluation of naloxone efficacy/safety, it might impact the size of the patients' population eligible for randomization, since it is hypothetised that the tested standardized protocol of AEDs withdrawal will reduce the frequency of SAEs including SGTCS in patients not usually suffering this seizure type. However, this impact on ENALEPSIE recruitment will be limited for several reasons : (i) According to SAVE cluster-randomised design, stratification, and overlap of participating centers with ENALEPSIE, only five or six ENALEPSIE centers should be randomised to receive the standardized protocol of AEDs withdrawal tested in SAVE. All other ENALEPSIE centers will continue using their current non-standardised practice of AEDs withdrawal. Secondly, the impact expected from SAVE intervention on the occurrence of SGTCS is also limited, with an estimation that the latter will decrease by 20%. Overall, SAVE might reduce the overall number of SGTCS by less than 9%. Our contingency plan, in case both studies are to be implemented simultaneously, will be to include two more participating adult French EMUs into ENALEPSIE.

**- According to the characteristics of ENALEPSIE and SAVE studies, French ethical rules and regulations allow simultaneous inclusion of patients in both studies.**

##### 6.2 INCLUSION CRITERIA

For inclusion:

- Adult patient (≥ 18 years) suffering from drug-resistant partial epilepsy.

- Patient undergoing long-term video-EEG or video-SEEG monitoring in one of the participating centre to record and characterize its seizure.

- Patient who gave its written informed consent to participate to the study,

- Patient affiliated to the French health care system

For randomization: Patient who suffers a secondary generalized tonic-clonic seizure during the long-term video-EEG or video-SEEG monitoring while being supervised by a nurse or a physician.

##### 6.3 EXCLUSION CRITERIA

- Age < 18 years.

- Patient that has already been randomized in this study

- Patient who is protected adult according to the terms of the French law
- Pregnant or breastfeeding women.

- Hypersensitivity to naloxone.History of severe heart disease (myocardial infarction, heart failure disorder, arrhythmia

severe hypertension)
- Ongoing opioïd treatment, including both pure agonists and partial agonists.
- Addiction to opioïds, heroin, or any similar substance.

- Participation in other drug clinical trial within the last two months

##### 6.4 EARLY WITHDRAWAL OF SUBJECTS

Criteria for removal from study

- Consent withdrawal

- Requirement of opioid treatment to control pain during video-EEG/video-SEEG monitoring

As detailed in section 6.3, punctual administration of opioïd treatment will be authorized, but will result in temporary ineligibility for randomization (12 hours). However, if pain management requires administration of opioid treatment during three consecutive days, the patient will be excluded from the study.

- Adverse event

Treatment will be discontinued or patient withdrawn in case of severe adverse event leading to significant risks for the patient. This will be appreciated by the investigator, according to the severity of the adverse event on the one hand and to the degree of certainty of the responsibility of the study treatment in the occurrence of adverse event on the other. The decision of unblinding will be decided on a case by case basis by the investigator in case of serious adverse event. In general way, unblinding procedure will be restricted to clinical situations in which knowledge of treatment allocation is likely to influence the management of the adverse event. The unblinding procedure is discussed in section 8.2.

Data collection and follow-up for withdrawn subjects

If a subject does not return for a scheduled visit, every effort should be made to contact the subject. In any circumstance, every effort should be made to document subject outcome, if possible. The investigator should inquire about the reason for withdrawal, request the subject(s) to return for a follow up visit, if applicable and follow-up with the subject regarding any unresolved adverse events. If the subject withdraws from the study, and also withdraws consent for disclosure of future information, no further evaluations should be performed, and no additional data should be collected. The sponsor may retain and continue to use any data collected before such withdrawal of consent.

#### 7 TREATMENTS

Patients will be randomized into one of the two study arms: naloxone or placebo.

##### 7.1 STUDY DRUGS

###### 7.1.1 Naloxone

Naloxone is a specific opioid antagonist, without partial agonist activity. Intravenous administration is approved in France since 1977 for the treatment of acute opioid overdose. In this indication, the recommended starting dose is 0.4 mg (Boyer, 2012). In acute opiate overdose, the delay between intravenous injection and awakening ranges from 30 seconds to 2 minutes and its apparent duration of action is 20 to 90 minutes (Boyer, 2012). No epilepsy-related alert has been reported for this product. Naloxone will not be use in its indication.

Naloxone will consist in dihydrous hydrochloride naloxone (0.4 mg/ml) packaged in 1 ml vials which can be intravenously administered to the patient without additional dilution. In order to respect the blinding, naloxone will be prepared by the pharmaceutical department of Edouard Herriot Hospital (Hospices Civils de Lyon).

###### 7.1.2 Placebo

Placebo will be isotonic sodium chloride which preparation in 1 ml vials will be centralized by the pharmaceutical department of Edouard Herriot Hospital to ensure its indistinguishability from naloxone.

###### 7.1.3 Preparation and disribution

Naloxone and placebo vials will be prepared by the pharmaceutical department of Edouard Herriot Hospital using aseptic technique to ensure the sterility of the prepared solution according to French good preparation practices.

Study treatments will be distributed by the pharmaceutical department of Edouard Herriot Hospital to the pharmacies of other investigator sites.

###### 7.1.4 Storage and dispensation

The study treatments will be stored at room temperature at the hospital pharmacies of investigator sites. Because of urgent need of treatment in case of occurrence of GTCS, one or more study treatments will be stored at room temperature in each EMU according to the expected number of randomizations of each center. New batches will be provided to the EMU by the pharmacy after each randomization. Dispensing will be made according to local regulations. Each pharmacy will keep dispensing records and will be in charge of the accountability of the study treatment.

Unused study batches will be destroyed following local procedures and the destruction will be documented in the drug accountability log. A certificate of destruction will be sent to the trial manager.

##### 7.2 DRUG ADMINISTRATION

###### 7.2.1 Method of assigning subjects to treatment group

The randomization lists will be centrally generated by the Clinical Research Unit of Lyon Hospital (Pôle IMER – Hospices Civils de Lyon). The pharmaceutical department of Edouard Herriot Hospital will prepare numbered vials according to the randomization lists.

Subjects who fulfill all inclusion/exclusion criteria will be included in the study and will receive a patient's number. If they develop GTCS, they will be randomized and will be assigned to a study arm (naloxone or placebo) according to the randomization procedure (see section 8.1).

###### 7.2.2 Administration

In case of occurrence of a supervised GTCS, a single dose of 1 ml of naloxone (i.e 0.4 mg) or placebo will be administered by the supervising nurse or physician, using direct intravenous injection with a 5 ml syringe through peripheral venous catheter of which all patients will be equipped throughout the video-EEG.

The evolution from a partial seizure to a GTCS being gradual, and the total duration of the seizure ranging from 2 to 3 minutes, the injection will be prepared during the course of the seizure. Given the assumptions about the role of endogenous opioids release in the spontaneous termination of seizures, naloxone will be administrated immediately after the end of the GTCS and not before. Specifically, treatment administration will be performed within the 2 minutes following the end of the GTCS.

#### 7.3 PRIOR AND CONCOMITANT THERAPY

Naloxone will not be allowed in patients with ongoing opioïd treatment, including both pure agonists and partial agonists. Therefore, patients with ongoing opioïd treatment will not be included. Use of extended-release formulation of opioid agonists will not be allowed during the study. Pain therapy in the included patients will be managed as following: (i) paracetamol will always be proposed as first-line therapy, (ii) if pain is not controlled with paracetamol, punctual administration of opioïd treatment will be authorized. However, this will result in temporary ineligibility for randomization during the 12 hours following opioid administration (excluding controlled-release formulations of morphine which are not used in these circumstances, elimination half-life of morphine and/or opioid agonists, including codein and tramadol, is about 4-8 hours). If pain management requires administration of opioid treatment during three consecutive days, the patient will be excluded from the study.

All antiepileptic drugs will be allowed during the study. Furthermore, inclusion in the study will not modify the management of antiepileptic drugs during the video-EEG monitoring. It is common practice reducing the dosage of antiepileptic drugs to promote a timely occurrence of seizures during video-EEG/video-SEEG. However, inclusion in the study will not modify the antiepileptic drugs withdrawal, the management of which will remain at the discretion of each center.

### 8 STUDY CONDUCT

This study will be conducted as outlined in the following.

#### 8.1 CLINICAL INVESTIGATIONS

Patients will be seen by study investigator at study inclusion (V0), which will correspond to the day immediately following the admission day in the EMU both for patients who undergo video-EEG and for those who undergo video-SEEG, and at the end of the monitoring (V2). Visits V0 and V2 will include a standard clinical evaluation, including the occurrence of adverse events. Patients will be randomized in case of occurrence of a GTCS during the video-EEG/video-SEEG monitoring (V1). All included patients will benefit from a phone visit (T1) by the CRA of the center 15 days after the end of the monitoring to retrieve late adverse events.

#### 8.2 VIDEO-EEG MONITORING

The modalities of the video-EEG monitoring will be consistent with the current practices and similar across the 8 centres. Patient preparation will include the installation of scalp electrodes for EEG recording and the installation of precordial electrodes for EKG recording. Similarly, the modalities of the video-SEEG monitoring will be consistent with the current practices and similar across the eight centres in which intracranial recordings are performed. In both video-EEG and video-SEEG, the standard preparation will also include installation of a digital photoplethysmographic sensor connected to a pulse oximeter to record SpO2. Thanks to a previous PHRC national (REPOMSE study), all participating centres indeed benefit from systematic recording of pulse oximetry coupled with video-EEG/video-SEEG. The systematic measurement of SpO2 provides additional security to video-EEG/video-SEEG mpnitoring, allowing early detection of rare apnea severe enough to be life-threatening. The reliability of the measurement of SpO2 during seizures has been validated by two studies which carried out this type of monitoring (Nashef et al., 1996; Bateman et al., 2008).

During video-EEG/video-SEEG, patients are continuously supervised by one to two specialized nurses during daytime in all partipating centres. This continuous supervision allows immediate intervention in case of occurrence of a seizure. In six centres, patients are supervised by one specialized nurse during the night whereas they are not supervised in the other centres.

#### 8.3 VISIT SCHEDULE

**Prior visit**
The patient is admitted in the EMU to start long-term video-EEG/video-SEEG monitoring for presurgical evaluation purpose
All inclusion/exclusion criteria will be checked
The study will be presented to the patient and an information letter will be given to him
The patient will have a time of reflection (one day) before accepting to participate to the study
Start of the video-EEG/video-SEEG monitoring according to the procedure detailed in section 7.2.

Long-term video-EEG/video-SEEG monitoring typically lasts between 7 and 15 days, and occasionnaly up to 21 days. Seizure rarely occurs during the first two days of the monitoring. As a matter of fact, progressive antiepileptic drugs withdrawal is usually required during the first week of the monitoring to promote the occurrence of seizure. In this context, the probability that patient develops GTCS on the admission day in the EMU before its inclusion in the study is very low.

**Visit v0±1day**
V0 will be performed the day immediately following the admission day in the EMU
We will firstly check that the patient meets all inclusion criteria and none of the exclusion criteria
The patient will sign the informed consent.
A complete clinical exam will be performed including
Retrieval of epilepsy characteristics, including the monthly seizure frequency over the past three months, the total number of GTCS during the same period
Retrieval of concomitant therapies
Physical/neurological exam
Vital signs
Weight/height
12-lead EKG
Installation of peripheral venous catheter
Continuation of the video-EEG/video-SEEG monitoring according to the procedure detailed in section 7.2.

**Period between V0 and V1**
Video-EEG/video-SEEG monitoring will be continued in accordance with the procedure detailed in section 7.2.
Functionality of the peripheral venous catheter will be checked twice a day every day (i.e in the early morning and in the early afternoon) using injection of 5 ml isotonic sodium chloride. A third verification of the functionality of the peripheral venous catheter will be performed at the beginning of the night in the patients who will be supervised during the night. The peripheral venous catheter will be changed if needed, according to the current pratices.
Patient's eligibility for randomization will be continuously assessed throughout the monitoring, specifically regarding the transient ineligibility for randomization in case of punctual use of opioids to control pain resistant to paracetamol.
Given the organization of some EMUs (see section 7.2), a GTCS might occur during a period without supervision. Non supervised GTCS will not result in randomization and the patient will remain eligible for randomization in case of occurrence of another GTCS during the monitoring.

**Visit v1**

In case of occurrence of a supervised generalised tonic-clonic seizure, eligible patients will be randomized to receive intravenous naloxone (0.4 mg) or placebo. Given the assumptions about the role of endogenous opioids release in the spontaneous termination of seizures, naloxone will be administered immediately after the end of the GTCS and not before. Specifically, treatment administration will be performed by the supervising nurse or physician within the 2 minutes following the end of the GTCS (see section 7.2.2).

The ability to administer the assigned treatment within this short post-ictal period will be sustained by the following organisation:

The study drug will be made readily available in a specific location of the EMU where will also be located the syringe needed for the intravenous injection, allowing immediate access to the treatment for the supervising nurse or physician. Accordingly, one or more study vials will be stored at room temperature in each EMU according to the expected number of randomizations of each center,

The randomization procedure, detailed in section 8.1, has been designed to facilitate this step. According to stratification by centre, each EMU will dispose from its own randomization list, which will be independant from the randomizations in the other participating centers. This will allow the pharmaceutical department of Edouard Herriot Hospital to prepare and to number the treatment vials at each EMU level. Thus, in each participating EMU, the randomized patients will be consecutively assigned to study treatment following ascending numerical order (i.e patient #1 will receive study vial #1, patient #2 study vial #2,…). In this context, the supervising nurse or physician will only have to use the first available study batch stored in the EMU, following ascending number.

The evolution from a partial seizure to a GTCS being gradual, and the total duration of the seizure ranging from 2 to 3 minutes, the injection will be prepared during the course of the seizure.

The pharmacological characteristics of naloxone allow its direct intravenous administration to the patient without additional dilution (see section 6.1.1). In this context, treatment administration will consist in direct injection through the peripheral venous catheter of the content of the assigned vial using a 5 ml syringe. As noted above, the functionality of the peripheral venous catheter would have been assessed twice a day every day in all patients (i.e in the early morning and in the early afternoon), with an additional verification at the beginning of the night in those who will benefit from nocturnal supervision.

Immediately after the administration of the treatment, the supervising nurse or physician will performed the following tasks:

Analysing respiratory movements to detect absence of chest expansion

Asking the patient to execute a verbal command (shake hand, answer a question, take a pencil…) regularly depending on the condition of the patient (between 1 minute and 5 minutes, to renew). The time at which verbal command is performed will be noted.

The time at which the first spontaneous movement of the patient is performed will be noted.

Heartbeat and blood pressure will be assessed every 20 minutes within 2 hours after the administration of treatment.

Sp02 monitoring:

Systematic SpO2 monitoring being part of current practices in all participating centres in order to optimize the safety of video-EEG/video-SEEG monitoring, all supervising nurses have been trained to monitor this parameter during the post-ictal period.

In patients who show Sp02 < 85% during > 2 minutes after the injection of the study drug, the nurse will administer 02 using high concentration breathing mask (15 L/min).

In patients who demonstrate persistent respiratory distress and oxgen desaturation despite 02 inhalation using high concentration breathing mask (15 L/min), a specific cardiorespiratory rescue process will be initiated:

Intensive care unit, as well as study investigator, will be immediatly alerted of life-threatning event.

02 using high concentration breathing mask (15 L/min) is maintened and material for cardiac resuscitation is ready for use, including semi-automatic cardiac defibrillator of which all participating EMU are equiped

Intensivist physician will come to the EMU to evaluate the patient and to decide whether or not the situation requires its immediate transfer to the Intensive Care Unit or any other resuscitation procedure

After recovery of consciousness, pain will be assessed with analogic visual scale
Occurrence of one or more GTCS during the two hours following treatment administration
Removal of peripheral venous catheter two hours after the administration of the treatment

**Period between V1 and V2**
Video-EEG/video-SEEG monitoring will be continued in accordance with the procedure detailed in section 7.2.
Occurrence of another supervised GTCS during the monitoring will not result in a second randomization
Adverse events will be retrieved throughout this period

**Visit v2**
After a maximum duration of 3 weeks, the video-EEG/video-SEEG monitoring will end. The total duration of monitoring will be variable across patients, this latter being defined by clinical parameters, specifically the occurrence of a suffisant number of seizures to allow conclusion about the localization of the seizure onset zone.
A complete clinical exam will be performed
A time schedule for the phone visit T1 will be determined with the patient
The patient will leave the EMU

**Phone visit t1 : 15 days ±7 days after V2**
The patient is contacted by phone by the CRA of the center 15 days after its discharge from the EMU to ensure the absence of serious adverse event

#### 8.4 CHANGES TO THE CONDUCT OF THE STUDY OR PROTOCOL

Any changes in the study protocol, such as changes in the study design, objectives or endpoints, inclusion and exclusion criteria, and/or procedures will be implemented only after the mutual agreement on the Investigator coordinator and Sponsor. All protocol changes must be documented in protocol amendments. Protocol amendments must be signed by the Investigator coordinator and submitted to and approved by the EC. Documentation of EC approval must be returned to Sponsor. Protocol amendments will be submitted to all applicable regulatory agencies.

#### 8.5 STUDY FLOW-CHART

| ***Study period*** | **Epilepsy Monitoring Unit** | | | | | | **T1** **15 days ± 7 days after V2** |
|---|---|---|---|---|---|---|---|
| | **Admission** | **V0** | **Between V0 and V1** **Video-EEG / Video-SEEG** | **V1** | **Between V1 and V2** **Video-EEG / Video-SEEG** | **V2** **End of monitoring** | |
| **Study day** | **- 1** | **0** | ***** | ***** | ****** | **Max 21**** | **Max 36***** |
| Presentation of the study and information letter | ✓ | | | | | | |

| | | | | | | | |
|---|---|---|---|---|---|---|---|
| Consent signed | | ✓ | | | | | |
| Inclusion/exclusion criteria | ✓ | ✓ | | | | | |
| Eligibility criteria for randomization | | ✓ | ✓ | ✓ | | | |
| Randomization | | | | ✓ | | | |
| Administration of the treatment | | | | ✓ | | | |
| Start of video-EEG/video-SEEG monitoring | ✓ | | | | | | |
| Sp02 monitoring | ✓ | ✓ | ✓ | ✓ | ✓ | | |
| Continuous video-EEG/video-SEEG monitoring | | ✓ | ✓ | ✓ | ✓ | | |
| Clinical evaluation | | ✓ | | | | ✓ | |
| Pain analogic visual scale | | | | ✓ | | | |
| Assessement of post-ictal chest movements | | | | ✓ | | | |
| Evaluation of adverse events | | ✓ | ✓ | ✓ | ✓ | ✓ | ✓ |
| End of video-EEG/video-SEEG monitoring | | | | | | ✓ | |

* = Occurrence of GTCS cannot be determined a priori. In this context, the day of randomization will be variable across patients
** = After a maximum duration of 3 weeks, the video-EEG/video-SEEG monitoring will end. The total duration of the monitoring will be variable across patients, this latter being defined by clinical parameters, specifically the occurrence of a suffisant number of seizures to allow conclusion about the localization of the seizure onset zone
*** = The phone visit T1 will always be performed **15 days ± 7 days after V2**.

### 9 TRIAL PROCEDURES

#### 9.1 RANDOMIZATION PROCEDURE

Randomization will be centralized, stratified by centre and balanced by block of patients. For each study center, randomization lists will be prepared by the Clinical Research Unit of Lyon Hospital (Pôle IMER – Hospices Civils de Lyon). The pharmaceutical department of Edouard Herriot Hospital (Hospices Civils de Lyon) will prepare numbered batches according to the randomization lists.

In case of occurrence of a GTCS, patient will be randomized in a 1:1 ratio and assigned to:
- Naloxone group,
- Placebo group.

Batch number will be registered in the patient's Case Report Form (CRF).

The following information will be sent to the coordination center and the hospital pharmacy of the investigator site via mail/fax:

- Patient's initials (first letter of name and first letter of surname),
- Patient's number (allocated during the inclusion step),
- Batch number.

#### 9.2 BLINDING AND UNBLINDING PROCEDURES

This is a double-blind study: the patient, the supervising nurse or physician and the investigator will not be aware of the nature of the administered treatment in order to avoid any follow-up or measure bias. Naloxone and placebo will be prepared in vials by the pharmaceutical department of Edouard Herriot Hospital (Hospices Civils de Lyon).

If case of occurrence of serious adverse event, the Centre Anti-Poison of Lyon will proceed to unblinding 24h/24 upon request of the coordinator investigator, the investigators or the sponsor. A detailed written procedure for unblinding will be provided to all persons involved

#### 9.3 DIGITAL DATA EVALUATION PROCEDURE

All digital data (video, EEG, respiration, SpO2) will be centralized and evaluated blind to other data by the coordinator investigator of the study who will not be involved in the video-EEG monitoring of the included patients.

All digital data, including that for SpO2 analyses, will correspond to segments of video-EEG/video-SEEG files including the seizure. The file should include 5 minutes prior to the seizure, 15 minutes after the start of the seizure, including the seizure itself, the onset and the end of which will be estimated from the video-EEG/video-SEEG data and annotated in the file, and a post-ictal period. Two different files will be created. The first one will only include only SpO2 and EKG data, EEG and video data being stored in the second file. Both files should not include any mention of the name or surname of the patient, apart from the first letters defining the patient's number in the study. All data will be stored on CD and sent to the coordinator of the study.

The same automatic and objective analysis of SpO2 data than the one already developed in the PHRC REPOMSE will be performed. This latter used a specific detection algorithm which allowed automatic detection and quantification of SpO2 and EKG variations.

Analysis of EEG data, and specifically evaluation of PGES will use the same criteria and methods than that proposed previously (Lhatoo et al., 2010). However, PEGS will only be assessed in patients undergoing scalp Video-EEG. Given the limited brain sampling of invasive recordings, we cannot discrimate postictal focal EEG suppression from generalized EEG suppression in patients undergoing video-SEEG. In addition, the cutoff value which would define absence of electroencephalographic activity with intracranial electrodes remains unknown

### 10 SAFETY CRITERIA

#### 10.1 DEFINITIONS

##### 10.1.1 Adverse Events

An Adverse Event (AE) is any untoward medical occurrence in a patient or clinical investigation patient administered a pharmaceutical product and which does not necessarily have to have a causal relationship with this treatment. An adverse event can therefore be any unfavourable and unintended sign (including an abnormal laboratory finding, for example), symptom, or disease temporally associated with the use of a medicinal product, whether or not considered related to the medicinal product.

##### 10.1.2 Serious Adverse Events

Will be considered as Serious Adverse Event (SAE) any untoward medical occurrence that at any dose:

- Results in death
- Is life-threatening
- Requires in-patient hospitalization or prolongation of existing hospitalization
- Results in persistent or significant disability/incapacity
- Is a congenital anomaly/birth defect
- Is a medically significant event such as:
  - Severe haemorrhage
  - Severe hemodynamic failure
  - All visceral failure: renal insufficiency necessitate dialysis, respiratory failure needed respiratory assistance, liver failure (bilirubin >10N), brain failure
  - Home parenteral nutrition more than 1 month
  - All infectious complications (intra-abdominal abscess)
  - Grade 4 thrombopenia (<25 000), Neutropenia (<500 PN), Transfusion (more than 4 RBCU)
  - Diarrhoea > 15 per day after 21 days
  - All adverse effect affecting life prognosis or leading to permanent or temporary serious incapacity

Medical and scientific judgment should be exercised in deciding whether expedited reporting is appropriated in situations, such as important medical events that may not be immediately life-threatening or result in death or hospitalization but may jeopardize the patient or may require intervention to prevent one of the outcomes listed in the definition above*.*
The term "*severe*" is a measure of intensity, thus a severe adverse event is not necessarily serious. For example, "*nausea of several hours*" duration may be severe but may not be clinically serious.

#### 10.2 INTENSITY

The intensity of the event will be graded according to the four-point system below:

| | |
|---|---|
| Mild (grade 1) | Discomfort noticed but no disruption of normal daily activity |
| Moderate (grade 2) | Discomfort sufficient to reduce or affect normal daily activity |
| Severe (grade 3) | Incapacitating with inability to work or perform normal daily activity |
| Life-threatening (grade 4) | Substantial risk of dying at time of event |
| Death (grade 5) | |

#### 10.3 OBLIGATIONS OF THE INVESTIGATOR

##### 10.3.1 Adverse Events reporting

All adverse events (AE) regardless of seriousness or relationship to Investigational Product that occurred after the informed consent up to 30 days after the last study drug administration are to be recorded in the AE pages of the Case Report Forms (CRF).
Whenever possible, symptoms should be grouped as a single syndrome or diagnosis. The investigator should specify the date of onset, intensity, action taken regarding trial medication, corrective therapy given, outcome of all adverse events and his opinion as to whether the adverse event can be related to the study drugs
All events that meet one or more criteria of seriousness will be reported as Serious Adverse Event.

General AE/SAE reporting rules:

- Any episode of any grade, related to a Serious Adverse Event must be reported as "Adverse Event" in the appropriate CRF pages;

- Planned hospital admissions or surgical procedures for an illness or disease which existed before the patient was enrolled in the study or before study drug was given are not to be considered SAEs unless the condition deteriorated in an unexpected manner during the study (eg surgery was performed earlier than planned).

##### 10.3.2 Serious Adverse Events reporting

All Serious Adverse Events occurred after the informed consent up to 30 days after the last study drug administration, whether or not ascribed to the study, must be reported on the Adverse Event page of the CRF.
A Serious Adverse Event that occurs after this time, including during the follow-up period, if considered related to the study medication, will be reported.
Serious Adverse Event will not be recorded after the start of a new out-of-study treatment or after disease progression

In a case of Serious Adverse event, the Investigator must immediately:
- Send (within 1 working day by fax) the SAE pages to

**ENALEPSIE SAFETY DESK**
**Hospices Civils de Lyon**
**FAX: + 00 33 (0) 4 72 11 51 90**

All SAE forms must be dated and signed by the responsible Investigator or one of his/her authorized staff Members.
- Whenever possible, symptoms should be grouped as a single syndrome or diagnosis. The investigator should specify the date of onset, intensity, action taken regarding trial medication, corrective therapy given, outcome of all adverse events and his opinion as to whether the adverse event can be related to the study drugs (unrelated or related).
- Attach the photocopy of all examinations carried out and the dates on which these examinations were performed. Care should be taken to ensure that the patient’s identity is protected and the patient’s identifiers in the Clinical study are properly mentioned on any copy of source document. For laboratory results, include the laboratory normal ranges.
- Follow up of any Serious Adverse Event that is fatal or life threatening should be provided within one calendar week.

##### 10.3.3 Follow up of Adverse Events and Serious Adverse events

Any SAEs should be monitored until they are resolved or are clearly determined to be due to a patient’s stable or chronic condition or underlying condition. Any additional information known after the event has been initially reported should be sent to the SAFETY DESK as soon as information becomes available. All adverse events must be documented and the outcome must be followed up until the return to normal or consolidation of the patient’s condition.

#### 10.4 OBLIGATIONS OF THE SPONSOR

During the course of the study, the Sponsor will report in an expedited manner all SAEs that are both unexpected and at least reasonably related to study drug, to the Health Authorities, Ethic Committees in each country in accordance with international and local regulations, and to the Investigators.
The expectedness of an adverse reaction will be determined by the Sponsor according to the Summary Product Characteristics of naloxone and isotonic sodium chloride respectively.
The sponsor will report all safety information from the trial in the Annual Safety Reports and notify it to the Health Authorities and Ethics Committees in accordance with with international and local regulations.

### 11 INVESTIGATORS RESPONSABILITIES

#### 11.1 Patient Informed Consent

A patient must provide written consent before undergoing any protocol-required assessments. Written informed consent in compliance with local regulatory authority will be obtained from each patient prior to entering the trial. It is responsibility of the Investigator to obtain such consent. Sample informed consent form (ICF) documents will be provided to each centre. The patient and the investigator will date and sign the ICF. The investigator shall provide a copy of the signed consent to the study patient; a copy shall be maintained in the investigator's study file.

#### 11.2 Protocol Adherence

Each Investigator must adhere to the protocol as detailed in this document.
Each Investigator will be responsible for allowing only those who have met protocol eligibility criteria to be randomized. Modifications to the protocol should not be made without agreement of the Investigators and Sponsor. Changes to the protocol will require written ethic committee approval/favorable opinion prior to implementation.
The sponsor will submit all protocol modifications to the appropriate regulatory authorities in accordance with the governing regulations.

#### 11.3 Monitoring/Audit

A representative of Sponsor or designee will visit the Investigator periodically for the purpose of monitoring the progress of this study in accordance with GCP regulations. It is the responsibility the Investigator to be present or available for consultation during such scheduled monitoring visits. During these routine visits, all data pertaining to a patient's participation in this clinical investigation must be made available to the monitor. On-site review of the CRFs for completeness and clarity, cross checking with sources documents, and reconciliation and clarification of administrative matters will be performed. An audit may be performed at any time during of after completion of the clinical study by the Sponsor or designee. All study-related documentation must be made available to the designated auditor(s).
Monitor designated by the Sponsor will ensure that:

- Storage times and conditions are acceptable, and that supplies are sufficient throughout the trial;
- Investigational products have been supplied only to subjects who are eligible to receive it and at the protocol specified dose;
- The receipt, use and return of the investigational products at the trial site are controlled and documented adequately.

In addition, a representative of the regulatory agency may choose to inspect a study centre at any time prior to, during or after completion of the clinical study. A Sponsor representative or designee will be available to assist in the preparation for such an inspection. All pertinent study data should be made available as requested to the regulatory authority for verification, audit, or inspection purposes.

#### 11.4 Criteria for premature discontinuation of the treatment

Circumstances that lead to premature withdrawal of a patient from the trial must be clearly reported by the investigator on the appropriate CRF page.
Patients can be withdrawn from the study under the following circumstances:

- Death;
- Intercurrent illness;
- Non compliance (including loss of patient to follow-up);
- Voluntary withdrawal;
- Failure to meet the eligibility criteria.

Patients are free to withdraw from the study at any time without prejudice to their treatment. When a patient decides to withdraw from the study, he should always be contacted in order to obtain information about the reason for withdrawal and to record any adverse events.
Every effort will be made to contact patients who fail to return for scheduled visits. A patient is considered lost to follow-up if no information has been obtained when the patient has completed the last clinical phase of the study. During this time there must be documented attempts to contact the patient either by phone or letter.

##### 11.5 PREMATURE CLOSURE OF THE STUDY

Study participation by individual sites or the entire study may be prematurely terminated, if in the opinion of the sponsor, there is sufficient reasonable cause. Any investigator who wants to discontinue his/her participation to the study must immediately inform the sponsor in writing of this decision.
Written notification documenting the reason for study termination will be provided to the investigator by the terminating party.
Examples of circumstances that may warrant termination include:

- Failure to enter patients at an acceptable rate;
- Insufficient adherence to protocol requirements;
- Insufficient complete and/or evaluable data;
- Frequency and/or unexpected severity of a toxicity;
- Unacceptable toxicity.

#### 12 STATISTICAL ANALYSIS

##### 12.1 DETERMINATION OF SAMPLE SIZE

Sample size was revised using a log rank test with a two-sided alternative hypothesis. For a significance level of 5% (two-tailed), assuming a hazard ratio of 2.414 calculated, i.e based on an effect size similar to the effect size of oxygenotherapy in a delay of 60 seconds (Rheims et al., 2019), 40 events should be observed to reject the null hypothesis in 80% of cases,. A proportion of 89% of patients being expected to recover SpO2 ≥ 90% 120 seconds after the end of the seizure and considering a proportion of unusauble records for technical reasons of 20%, at least 54 patients should be randomized.

According to the REPOMSE study (Rheims et al., 2019), which included 1069 patients with similar clinical characteritics in the same epilepsy monitoring units, about 13% of patients with drug-resistant focal epilepsy who undergo long-term video-EEG monitoring develop at least one focal secondary generalized tonic-clonic seizure. Considering that some patients might not be elibiligle for randomisation because of contextual issues, including nocturnal occurrence of the seizure when the staff is reduced, we assume that about 10% of patients with drug-resistant focal epilepsy who undergo long-term video-EEG monitoring will be eligible for randomisation, a total of 554 patients will be included in the study.

##### 12.2 STATISTICAL ANALYSIS PLAN

The information presented below constitutes the basis for the statistical analysis plan for this study. This plan may be revised over the duration of the study in order to take into account any amendments to the clinical trial protocol, or adapt to any unforeseen difficulties in carrying out the study or with the data which impact the planned analysis. This revision will be based on an examination of the work and the data, and a final plan will be established prior to the cut-off point for finalizing the database. The statistical analysis will be performed independently by the investigators in order to ensure entirely objective results (Clinical Research Unit, Pôle IMER – Hospices Civils de Lyon).

###### 12.2.1 Population analysis

###### 12.2.1.1 Population “inclusion”

This population will comprise all included patients.

###### 12.2.1.2 Population “efficacy intention to treat”

This population will comprise all randomized patients

###### 12.2.1.3 Population “efficacy per-protocol”

This population will comprise all randomized patients having no major protocol deviation such as patients in whom the onset of intravenous injection of the study drug was within the two minutes following the end of the seizure (whereas those in whom the study drug was administered > 120 seconds after the end of the seizure will be excluded).

###### 12.2.1.4 Population “safety”

This population will comprise all patients who received naloxone or placebo injection.

##### 12.2.2 Statistical methods

###### 12.2.2.1 Population

A detailed diagram describing all populations will be presented. Patient flow with inclusions, exclusions and total numbers randomized to each arm will also be presented

###### 12.2.2.2 Protocol deviations

Any protocol deviation that may have an impact on the study results will be listed

###### 12.2.2.3 Background and demographic characteristics

Baseline patient characteristics will be summarized using descriptive statistics (number, mean, standard deviation, median, minimum and maximum for quantitative measures and numbers and percentages for qualitative measures).

###### 12.2.2.4 Analysis of the primary outcome measure

This analysis will be performed on the population "efficacy intention to treat". In order to keep patients whose oxygen saturation (SpO2) is ≥ 90% at the end of the seizure in this analysis, the null delay between the end of the seizure and recovery of oxygen saturation will be replaced by a non null value. This value will be smaller than the value observed for the first event whatever the arm in order to keep the rank of events. Kaplan-Meier survival analysis will be used to analyse the time to recovery of SpO2 ≥ 90%. If no censure is observed (i.e. no death) a Wilcoxon test will be used to compare the delays to saturation.

In case of important variability of the delay of injection after the end of the GCS, a Cox proportional hazards model will be used to adjust the estimation of time to saturation on time to injection. The level of significance for the arm effect will be 0.05.

This analysis will also be performed on the population "efficacy per-protocol".

###### 12.2.2.5 Analysis of the secondary outcome measures

This analysis will be performed on the population "efficacy" (unless otherwise specified).

secondary outcomes previously defined as proportions will be presented in each group (naloxone and placebo) and will be compared using Fisher exact test.

- secondary outcomes previously defined as delays and durations will be presented as mean (standard deviation) and median (minimum and maximum) in each group (naloxone and placebo) and will be compared between the two groups using a Wilcoxon-Mann-Whitney test. In case of censoring, a Kaplan meier estimation and a log rank test will be used.

- A listing of adverse events will be presented on the population "security". The percentages of occurrence of these adverse events (serious and non-serious) will be calculated in each group (naloxone and placebo). These percentages will be compared between the two groups using Fisher exact test.

- Pain evaluated by a visual analog scale, immediately after the recovery of consciousness following the postictal coma, will be presented as mean (standard deviation) and median (minimum and maximum) in each group (naloxone and placebo). These values will be compared between the two groups using a Wilcoxon-Mann-Whitney test.

### 13 QUALITY CONTROL

#### 13.1 RESPONSIBILITIES OF THE INVESTIGATORS

The investigator(s) undertake(s) to perform the study in accordance with Good Clinical Practice and specifically either good clinical practice for trials on medicinal products in the European Community.

The Investigators are required to ensure compliance with respect to the investigational drug schedule, visit schedule required by the protocol and all study procedures provided by the Sponsor. The investigators agree to provide reliable data and all information requested in the Case Report Form in an accurate and legible manner according to the instructions provided and to ensure direct access to source documents to Sponsor representatives.

The Investigator may appoint such other individual as he/she may deem appropriate as sub-Investigators to assist in the conduct of the study in accordance with the protocol. All sub-Investigators shall be appointed and listed in a timely manner. The sub-Investigator will be supervised by and work under the responsibility of the Investigator.

Patient compliance to the study treatment is the investigator's responsibility and will be checked during site monitoring visits by a representative of the sponsor.

The investigators agree to accept quality assurance audits performed by persons authorized by the promoter as well as inspections by the Competent Authorities. All data, all documents and reports may be subject to audits and regulatory inspections can be opposed without medical confidentiality.

##### 13.2 RESPONSIBILITIES OF THE SPONSOR

The sponsor of this study has responsibilities to Health Authorities to take all reasonable steps to ensure the proper conduct of the study as regards ethics, protocol adherence, integrity and validity of the data recorded on the case report forms.

A Clinical Research Associate (CRA) by the sponsor will ensure the successful completion of the study, the collection of data generated by writing, documentation, recording and report, in accordance with the Standard Operating Procedures implemented within the DRCI Hospices Civils de Lyon and in accordance with Good Clinical Practice and the legislative regulatory.

During monitoring visits, the following points will be scrutinized with the investigator: patient informed consent, patient recruitment and follow-up, study drug allocation, patient compliance to the study treatment, study treatment accountability, concomitant therapy use, adverse event documentation and reporting, and quality of data.

After all visits a monitoring report will be written and sent to the investigator visited and the coordination structure of the research. Investigators agree to accept quality assurance audits performed by persons authorized by the sponsor as well as inspections by the Competent Authorities. All data, all documents and reports may be subject to audits and regulatory inspections can be opposed without medical confidentiality

##### 13.3 STUDY DRUG MONITORING

Accountability for the study drug at the clinical site is the responsibility of the investigator. The investigator will ensure that the study drug is used only in accordance with this protocol. Where allowed, the investigator may choose to assign some of the drug accountability responsibilities to a pharmacist or other appropriate individual. Drug accountability records indicating the drug's delivery date to the site, inventory at the site, use by each patient, or disposal of the drug will be maintained by the clinical site. These records will adequately document that the patients were provided the doses as specified in the protocol. The sponsor will review drug accountability at the site on an ongoing basis during monitoring visits.

#### 14 DATA QUALITY ASSURANCE

##### 14.1 SOURCE DOCUMENT REQUIREMENTS

According to the guidelines for Good Clinical Practices, the study monitor has to check the case report form entries against the source documents. The Informed Consent Form will include a statement by which the patients allow the Sponsor's duly authorized personnel (trial monitoring team) to have direct access to original medical records which supports data on the electronic Case Report Forms (e.g. patient's medical file, appointment books, original laboratory records, etc.). These personnel, bound by professional secrecy, will not disclose any personal identity or personal medical information (according to confidentiality rules).

#### 14.2 Case Report Forms

Copies of pertinent records in connection with the study, including all source documents, will be made available to the Sponsor or its designee on request with due precaution towards protecting the privacy of the patient.

Case report forms (CRF) will be filled out legibly and completely. Unless explicitly directed, blank data fields are not acceptable. Any erroneous entries made on the CRFs will be crossed out with a single line, initialed and dated, and the correct entry, if appropriate, will be recorded. Writing in the margins of the CRFs is not permitted. The original CRFs will be provided to the Sponsor or its designee. A copy of the CRFs will be maintained in the Investigator’s site file. Illegible or incomplete entries or entries needing additional explanation will be returned or queried to the Investigator or designated representative for clarification. Erroneous values and/ or text must not be obliterated.

The report forms will include only the necessary data analysis for publication. Other patient data necessary for its monitoring outside of the study will be collected from the medical record.

This electronic case report forms will be established in each center with an internet support data collection. A help document for the use of this tool will be provided to investigators. Filling specifications observation via internet by the investigator allows the coordination center of the study quickly view and distance data.

In addition, during their seizures, these data are immediately checked by consistency checks. In this respect, it must validate any change in value in the CRF. These changes are subject to audit trail. Justification can be optionally integrated comment. Paper printing will be required at the end of study, authenticated (signed and dated) by the investigator. A copy of the authenticated to the developer shall be archived by the investigator.

#### 14.3 Archiving clinical trial files

The investigator shall maintain the essential clinical study documents (including source documents, clinical drug-disposition records, signed subject Information Consent Forms, AE reports, and other regulatory documents) as required by the applicable regulatory requirements. The investigator should take adequate measures to prevent accidental or premature destruction of these documents. In the event of accidental destruction, the investigator must notify Sponsor immediately. The following essential clinical study documents must be maintained: under the responsibility of the coordinating investigator and associate investigator at each center for 15 years :

- Signed informed consent documents for all subjects;
- Subject identification code list, screening log (if applicable), and enrollment log;
- Record of all communication between the investigator and ethics committee;
- Composition of the ethics committee or other applicable statement;
- Record of all communications between the investigator and the sponsor
- List of sub investigators and other appropriately qualified persons to whom the Principal Investigator has delegated significant trial-related duties, together with their roles in the study and their signatures;
- Copies of CRFs pages and of documentation of corrections for all subjects;
- Drug-accountability records;
- All other source documents (i.e., subject records, hospital records, laboratory records, etc.);
- All other documents as listed in Section 8 of the consolidated guideline on GCP (Essential Documents for the Conduct of a Clinical Trial). Essential clinical study documents shall be retained for at least 15 years following the date of the end of the study.

These documents shall be retained for a longer period, however, if required by additional applicable regulatory requirements or by an agreement with the sponsor. The investigator must, therefore, obtain approval in writing from the sponsor prior to destruction of any records.

The investigator shall notify the sponsor to any change in the location or status of any essential, clinical-study documents. The sponsor shall be responsible for informing the investigator when these documents no longer need to be retained.

##### 14.4 CNIL

This study is part of the "Methodology Reference" (MR-001) Article 54 paragraph 5 of the Law No. 78-17 of 6 January 1978 relating to data, files and freedoms. This change was approved by decision of 5 January 2006. The sponsor, signed a commitment to comply with the "Methodology of Reference."

#### 15 ETHICS, REGULATORY & LEGAL CONSIDERATION

##### 15.1 RISK/BENEFIT RATIO

The main benefit of the study is to show that naloxone can reduce postictal central respiratory dysfunction in patients with epilepsy. Naloxone would thus be the first and only therapeutic approach which could be immediately delivered to reverse postictal apnea, especially during long-term video-EEG monitoring. In the absence of alternative therapy, there will be no loss of oppoturnity for patients assigned to placebo. Given the pathophysiological link between the occurrence of central apnea in the aftermath of GTCS and the risk of SUDEP, for which no preventive treatment is currently available, the demonstration that an opioid antagonist can effectively reduce postictal apnea would pave the way for an assessment of a preventive chronic therapy targeting the same pathophysiological pathway using naltrexone, an orally administered opioid antagonist (see section 5).

The risks of the study are limited. The main issues are related to the potential physiological effect of endogenous opioids in the postictal period, specifically their antiepileptic effect and their antalgic effect. Antagonising a mechanism thought to participate to seizure termination could theoretically aggravate seizures. It should however be noted that no epilepsy-related alert has been reported for naloxone or naltrexone (Ray et al., 2010; Boyer, 2012), even in patients with chronic alcoholism who however demonstrate high risk of seizures (Samokhvalov et al., 2010). Some patients can develop pain during the course of the seizure or in the postictal period (Bell et al., 1997; Ekstein and Schachter, 2010). However, given the loss of consciousness which occur during GTCS, patients do not usually experience pain during seizure or in its immediate aftermath (Bell et al., 1997). Furthermore, postictal pain, including aches or postictal headache, usually do not require analgesic treatment or can be controlled with non-opioid analgesics (Ekstein and Schachter, 2010). In this context, and taking also into account the naloxone action duration (20 to 90 minutes (Boyer, 2012)), the risk of naloxone-induced increase of postictal pain appears limited.

Overall, we consider that the risk/benefit ratio appears favorable.

##### 15.2 ETHICAL CONDUCT OF THE STUDY

This study will be conducted in accordance with

- The Declaration of Helsinki on Ethical Principles for Medical Research Involving Human Subjects, adopted by the General Assembly of the World Medical Association (Seoul 2008, revised);
- The International Conference on Harmonisation Guidance on Good Clinical Practice (Topic E6) (CPMP/ICH/135/95);

- The European Clinical Trials Directive 2001/20/EC that provide greater protection to subjects participating in clinical trials, ensure quality of conduct and harmonise regulation and conduct of clinical trials throughout Europe;
- The law N°78-17 of 06/01/1978, modified in relation to computer science, to databases and to data collection;
- The law 2004-806 of August 9, 2004 defines the scope of the law as all biomedical research involving human beings, with the aim of increasing biological or medical knowledge;
- Bioethics law N°2004-800 of August 6 2004.

##### 15.3 REGULATORY AUTHORITY APPROVALS/AUTHORIZATIONS

Regulatory Authority approvals/authorizations/notifications, where required, must be in place and fully documented prior to study start. In France, the Sponsor will submit the study protocol for authorisation to the competent authority "Agence Française de Sécurité du Médicament et des produits de santé (ANSM)".

##### 15.4 SUBJECT INFORMATION AND CONSENT

It is the responsibility of the investigator to obtain informed consent in compliance with national requirements from each patient prior to entering the trial or, where relevant, prior to evaluating the patient's suitability for the study.

It must be made completely and unambiguously clear to each patient that they are free to refuse to participate in the study, or that they can withdraw their consent at any time and for any reason, without incurring any penalty or withholding of treatment on the part of the investigator.

The informed consent document used by the investigator for obtaining patient's informed consent must be reviewed and approved by the sponsor prior to Ethical Committee submission (Comité de Protection des Personnes Sud-Est II).

##### 15.5 EXCLUSION PERIOD

After inclusion, no simultaneous participation to other interventional clinical research will be authorized during ENALEPSIE study. At the end of ENALEPSIE study, no exclusion period is defined.

#### 16 ADMINISTRATIVE PROCEDURES

##### 16.1 RESEARCH BUDGET

This project is financially supported under the call for projects of the PHRC 2013

##### 16.2 INSURANCE

The sponsor certifies having taken out a liability insurance policy which covers the investigator and his co-workers and which is in accordance with the local laws and requirements (SHAM - 18 rue Edouard Rochet - 69372 Lyon Cedex 08 - contract number 144.244). Specific statements will be contained in appendix where is needed. A certificate of insurance will be provided to the investigator.

##### 16.3 INSPECTIONS BY REGULATORY AUTHORITIES

For the purpose of ensuring compliance with good clinical practice and regulatory agency guidelines it may be necessary to conduct a site audit or an inspection.

By signing this protocol, the investigator agrees to allow the sponsor and its representative, and drug regulatory agencies to have direct access to his study records for review. These personnel, bound by professional secrecy, will not disclose any personal identity or personal medical information.

These audits involve review of source documents supporting the adequacy and accuracy of data gathered in CRF, review of documentation required to be maintained, and checks on drug accountability.

The sponsor will in all cases help the investigator prepare for an inspection by any regulatory authority.

##### 16.4 PROTOCOL AMENDMENTS

No changes or amendments to this protocol may be made by the investigator or by the sponsor after the protocol has been agreed to and signed by both parties unless such change(s) or amendment(s) have been fully discussed and agreed upon by the investigator and the sponsor.

Any change agreed upon will be recorded in writing, the written amendment will be signed by the investigator and by the sponsor and the signed amendment will be appended to this protocol.

Approval/Autorisation of amendments by Health authorities (ANSM/Ethical Committee (Comité de Protection des Personnes Sud-Est II) is required prior to their implementation, unless there are overriding safety reasons.

#### 17 DATA AND SAFETY MONITORING BOARD (DSMB)

An independent Data and Safety Monitoring Board (DSMB) external to the trial investigators has been established specifically to monitor data throughout the life of a study to determine if it is appropriate, from both the scientific and ethical standpoint, to continue the study as planned. The DSMB will meet to review SAEs and propose to continue or stop the study. The DSMB is made up of three experts in epilepsy, pharmacology and methodology in clinical research.

Their role is to protect the interests of patients in the study and of those still to be entered by review of accumulating safety and tolerability data generated in the study. The sponsor will provide the DSMB with data by treatment arm to include demographic data, number of patients in study, duration of study drug exposure, AEs, SAEs, laboratory data and response/relapse. Based upon their review the DSMB will advise the Sponsor and the Steering Committee of any actions that should be taken in the study.

A meeting with the members of the DSMB will be held at least once a year or when required to discuss continuation of the study according to the occurrence of potential side effects.

#### 18 STUDY STERRING COMMITTEE

A steering Committee has been established to provide scientific guidance for this clinical study. This committee is comprised of experts who will assist the sponsor in revolving issues and/or questions encountered during the conduct of the study, and will consult with the sponsor regarding changes to the protocol as necessary. This committee is composed of the following experts:

- Pr Sylvain RHEIMS, Service de Neurologie Fonctionnelle et d'Epileptologie, Hospices Civils de Lyon
- Pr Fabrice BARTOLOMEI, Centre Hospitalier Universitaire de la Timone, Marseille
- Dr Arnaud BIRABEN, Centre Hospitalier Régional et Universitaire Pontchaillou, Rennes
- Dr Philippe CONVERS, CHU Hôpital Nord, Saint-Etienne
- Dr Arielle CRESPEL, Centre Hospitalier Universitaire Gui de Chauliac, Montpellier
- Pr Bertrand DE TOFFOL, Centre Hospitalier Universitaire Bretonneau, Tours

- Pr Philippe DERAMBURE, Centre Hospitalier Régional et Universitaire Roger Salengro, Lille
- Pr Edouard HIRSCH, Hôpitaux Universitaires de Strasbourg, Strasbourg
- Pr Philippe KAHANE, Centre Hospitalier Universitaire Michallon, Grenoble
- Pr Louis MAILLARD, Centre Hospitalier Universitaire de Nancy, Nancy
- Dr Véronique MICHEL, Centre Hospitalier Universitaire Pellegrin Tripode, Bordeaux
- Pr Vincent NAVARRO, Centre Hospitalier Universitaire de la Pitié-Salpêtrière, Paris
- Dr Jérôme PETIT, Centre de Lutte contre l'Épilepsie, Établissement de la Teppe, Tain l'Hermitage
- Dr Luc VALTON, Hôpital Rangueil, Toulouse
- Pr François CHAPUIS, Unité Recherche Clinique, Pôle Information Médicale, Evaluation et Recherche, Hospices Civils de Lyon, Lyon.

### 19 DURATION OF THE STUDY AND SCHEDULE

- Regulatory procedures: January 2014 to November 2014
- Patient recruitment (66 months): June 2015 to December 2020
- Last Patient Last Visit: January 2021
- Cut-off for the final clinical database: Second quarter 2021
- Analysis of final data: Third quarter 2021
- Interpretation and publication of results: Third quarter 2021

### 20 PUBLICATION OF TRIAL RESULTS

All the data collected during this study are the property of the sponsor and may not be communicated to third parties in any event without the written agreement of the study coordinating investigator.

Any publication or communication (oral or written) will be decided by common agreement among the investigators and will comply with international recommendations: "Uniform Requirements for Manuscripts Submitted to Biomedical Journals" (http://www.cma.ca/publications/mwc/uniform.htm).

Individuals who participated in the development of the study protocol, its progress and in the writing up of results will be the first signatories. The first author is one who take the initiative of the manuscript and who will be the main editor. All the investigators who included or monitored patients as a part of this research as well as the other collaborators involved will also be mentioned. In every publication, the Hospices Civils de Lyon will be named as sponsor and funding under the PHRC will figure explicitly.

ENALEPSIE study will be registered into a public clinical trials database (http://clinicaltrials.gov).
