## Supplementary material for "Efficacy of Naloxone in reducing hypoxemia and duration of immobility following focal to bilateral tonic-clonic seizures": ENALEPSY Statistical Plan

### ENALEPSIE

#### Efficacy of Naloxone in reducing postictal central respiratory dysfunction in patients with epilepsy.

Statistical analysis plan

Version 4: revision after unblinding

Date: 10/05/2021

Author: Catherine Mercier

Validation: Pascal Roy, Sylvain Rheims

**Sponsor:** HCL

**Coordinator investigator:** Dr Sylvain Rheims

**Coordination and Data Centre: Mathilde LECLERCQ** (data management with MedSharing)

**Statisticians:** Dr. Catherine Mercier, Pr. Pascal Roy

| History of previous versions | | |
|---|---|---|
| **Version** | **Date** | **Reason for the update** |
| V1.0 | 2020 09 28 | Original Version |
| V2.0 | 2020 11 18 | Amendment Primary endpoint and sample size Protocol V9 |
| V3.0 | 2021 02 26 | Amendment Primary endpoint Protocol V10 |
| V4.0 | 2021 05 10 | Deviations to randomization |

### SIGNATURE PAGE

**Trial biostatisticians**

Name: Date:

Pr Pascal ROY 04/12/2023

Name: Date:

Fatima CHORFA 04/12/2023

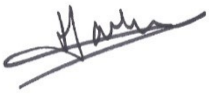

**Approved by:**

**Coordinating and leading principal investigator**

Name: Date:

Pr Sylvain RHEIMS

### Introduction

This version is a revision of the statistical analysis plan aim to take into account the major deviations to treatment allocation planned by randomisation. It replace the version validated before the closure of the data base and before the unblinding.

### 1 Objectives

#### 1.1 Primary objective

The main objective of the study is to evaluate the efficacy of 0.4 mg intravenous naloxone, versus placebo, administered in the immediate aftermath of a generalized tonic–clonic seizure (GTCS), in reducing the severity of the postictal central respiratory dysfunction occurring after the end of the seizure, as measured by pulse oximetry.

#### 1.2 Secondary objectives

- Assess the impact of naloxone on respiratory parameters such as the frequency of apneas (> 10 seconds) occurring after the end of the seizure.
- Assess the impact of naloxone on 02 administration requirement and on cardiorespiratory rescue procedure requirement after the end of the seizure.
- Assess the impact of naloxone on the postictal generalized EEG suppression, the duration of which is correlated with the risk of SUDEP, and the severity of which could also result from a seizure-related release of endogenous opioids.
- Assess the impact of naloxone on the duration of the postictal coma following a GTCS, and the time required by the patient to recover preictal clinical condition. In the future, this clinical benefit might be enough significant to justify the systematic use of naloxone after GTCS in inpatients
- Assess the frequency and severity of adverse events related to the treatment with naloxone, such as increased posictal pain and/or early recurrence of GCTS.

### 2 Trial design

DOUBLE-BLIND RANDOMIZED PLACEBO-CONTROLLED TRIAL

In case of occurrence of a generalised tonic-clonic seizure, patients will be randomized (1:1) into two groups:
Experimental group: patients with drug-resistant partial seizures who suffer a secondary generalised tonic-clonic seizure during long-term video-EEG monitoring and in whom naloxone is administered intravenously immediately after the end of the seizure.

Control group: patients with drug-resistant partial seizures who suffer a secondary generalised tonic-clonic seizure during long-term video-EEG monitoring and in whom placebo is administered intravenously immediately after the end of the seizure.

#### 2.1 Randomisation and blinding

Randomization is centralized, stratified by centre and balanced by block of patients (prepared by the Clinical Research Unit of Pôle IMER – Hospices Civils de Lyon). In case of occurrence of a GTCS, patient are randomized in a 1:1 ratio and assigned to:

- Naloxone group,
- Placebo group.

Batch number is registered in the patient's Case Report Form (CRF).
This is a double-blind study. Naloxone and placebo are prepared in vials by the pharmaceutical department of Edouard Herriot Hospital (Hospices Civils de Lyon). If case of occurrence of serious adverse event, the Centre Anti-Poison of Lyon will proceed to unblinding 24h/24 upon request of the coordinator investigator, the investigators or the sponsor.

#### 2.2 Time schedule

| *Study period* | Epilepsy Monitoring Unit | | | | | | T1 |
|---|---|---|---|---|---|---|---|
| | Admission | V0 | Between V0 and V1<br><br>Video-EEG/Video-SEEG | V1 | Between V1 and V2<br><br>Video-EEG/Video-SEEG | V2 | |
| **Study day** | **- 1** | **0** | ***** | ***** | ****** | **Max 21**** | **Max 36***** |
| Presentation of the study and information letter | ✓ | | | | | | |
| Consent signed | | ✓ | | | | | |
| Inclusion/exclusion criteria | ✓ | ✓ | | | | | |
| Eligibility criteria for randomization | | ✓ | ✓ | ✓ | | | |
| Randomization | | | | ✓ | | | |
| Administration of the treatment | | | | ✓ | | | |
| Start of video-EEG/video-SEEG monitoring | ✓ | | | | | | |
| Sp02 monitoring | ✓ | ✓ | ✓ | ✓ | ✓ | | |
| Continuous video-EEG/video-SEEG monitoring | | ✓ | ✓ | ✓ | ✓ | | |
| Clinical evaluation | | ✓ | | | | ✓ | |
| Pain analogic visual scale | | | | ✓ | | | |
| Assessement of post-ictal chest movements | | | | ✓ | | | |
| Evaluation of adverse events | | ✓ | ✓ | ✓ | ✓ | ✓ | ✓ |
| End of video-EEG/video-SEEG monitoring | | | | | | ✓ | |

* = Occurrence of GTCS cannot be determined a priori. In this context, the day of randomization will be variable across patients
** = After a maximum duration of 3 weeks, the video-EEG/video-SEEG monitoring will end. The total duration of the monitoring will be variable across patients, this latter being defined by clinical parameters, specifically the occurrence of a suffisant number of seizures to allow conclusion about the localization of the seizure onset zone
*** = The phone visit T1 will always be performed 15 days after the end of the monitoring.

#### 2.3 Endpoints

##### 2.3.1 Primary Endpoint

Delay between the end of the GTCS and recovery of oxygen saturation (SpO2) ≥ 90%.

##### 2.3.2 Secondary Endpoints

- Other respiratory parameters
  - Delay between onset of intravenous injection of naloxone and recovery of oxygen saturation (SpO2) ≥ 90%
  - Proportion of patients whose SpO2 is <70%, between 70 and 80%, between 80 and 85%, between 85 and 90% during at least 5 seconds (required to eliminate artifactual changes of SpO2), 30 seconds 1 minute, 2 minutes, 3 minutes, 4 minutes and 5 minutes after the end of GTCS of the study drug in the immediate aftermath of a GTCS.
  - Desaturation nadir in the immediate aftermath of a GTCS
  - Proportion of patients who show postictal apnea, defined as the absence of chest expansion during a period > 10 seconds between 30 seconds and 5 minutes after onset of intravenous injection of the study drug in the immediate aftermath of a GTCS. Apnea will be assessed by a combination of: 1) standardised observation and report from the nurse or physician who performed the postictal intravenous injection, and 2) video recording of the event. **Apnea is not reliable and will not be analysed**.
  - Proportion of patients in whom 02 administration is required within the ten minutes following the end of a GTCS.
  - Proportion of patients in whom cardiorespiratory rescue procedure is required within the ten minutes following the end of a GTCS
  - Total duration of the postictal generalized EEG suppression, defined as lack of detectable EEG activity >10 mV in amplitude on all leads. This ouctome will only be assessed in patients undergoing scalp Video-EEG
  - Total duration of the postictal coma, defined as the delay between the end of the seizure and the recovery of consciousness assessed by the ability to meet one single verbal command (handshake).
  - Total duration of the postictal immobility, defined as the delay between the end of the seizure and the first spontaneous movement of the patient, as assessed on the video recording.

This population will comprise all included patients.

##### 4.1.2 Population “eligible to randomization”

This population will comprise all patients eligible for randomisation.

##### 4.1.3 Population “randomized”

This population will comprise all randomized patients

##### 4.1.4 Populations “modified intention to treat”

This population (mITT) will comprise all randomized patients with GTCS. Patient who did not have GTCS being out of the main objective, they will be excluded from the analysis. Patients without any Spo2 data will be also excluded for the primary endpoint analysis.

##### 4.1.5 Population “per-protocol”

This population (PP) will comprise all randomized patients (mITT for the primary endpoint) having no major protocol deviation.

##### 4.1.6 Population “safety”

This population will comprise all patients who received naloxone or placebo injection = population randomized patients (all have received the injection).

#### 4.2 Protocol deviations

The most important deviation to the protocol which justify this SAP revision was the discrepancy between allocated arm and the received treatment, i.e. 20 patients out of 49 did not received the treatment planned by the randomization (40.8%):

| Allocated arm | Received treatment | | |
|---|---|---|---|
| Frequency | naloxone | placebo | Total |
| naloxone | 21 | **3** | 24 |
| placebo | **17** | 8 | 25 |
| Total | 38 | 11 | 49 |

- Patients in whom the study drug administered was not conform to the arm allocated by randomisation

| ID_CENTRE | ID_DOSSIER | Order | Randomized | Received | Allocated arm |
|---|---|---|---|---|---|
| 001 | 00258 | 1 | 14/09/2016 | naloxone | placebo |
| 006 | 00444 | 1 | 02/02/2017 | naloxone | placebo |
| 006 | 00804 | 2 | 18/01/2018 | naloxone | placebo |
| 007 | 00376 | 3 | 22/12/2016 | naloxone | placebo |
| 007 | 00377 | 2 | 13/12/2016 | naloxone | placebo |
| 009 | 00041 | 2 | 04/02/2016 | naloxone | placebo |
| 009 | 01557 | 4 | 12/06/2020 | naloxone | placebo |
| 010 | 00029 | 3 | 04/01/2016 | placebo | naloxone |
| 010 | 00044 | 4 | 11/02/2016 | naloxone | placebo |
| 010 | 00059 | 5 | 10/03/2016 | placebo | naloxone |
| 010 | 00170 | 7 | 22/06/2016 | naloxone | placebo |
| 010 | 00564 | 9 | 08/06/2017 | naloxone | placebo |
| 010 | 00711 | 12 | 11/10/2017 | naloxone | placebo |
| 010 | 01229 | 16 | 19/04/2019 | naloxone | placebo |

| ID_CENTRE | ID_DOSSIER | Order | Randomized | Received | Allocated arm |
|---|---|---|---|---|---|
| 010 | 01308 | 18 | 22/07/2019 | naloxone | placebo |
| 010 | 01565 | 22 | 22/06/2020 | naloxone | placebo |
| 013 | 00489 | 5 | 28/03/2017 | naloxone | placebo |
| 013 | 01409 | 7 | 04/11/2019 | placebo | naloxone |
| 013 | 01488 | 8 | 30/01/2020 | naloxone | placebo |
| 013 | 01515 | 9 | 17/02/2020 | naloxone | placebo |
| N = 20 | | | | | |

The other deviations to protocol already seen in blind review were the following:

- Patients wrongly randomised because the type of seizure was not GTCS (n=3)

| Subjid | VALIDE_COMPLET | Allocated arm | Received |
|---|---|---|---|
| 09-0047 | CRISE FOCALE | naloxone | naloxone |
| 10-0004 | CRISE FOCALE | naloxone | placebo |
| 10-0155 | CRISE FOCALE | naloxone | naloxone |

- Patients in whom the study drug was administered > 120 seconds after the end of the seizure (n=6)

| Subject ID | Allocated arm | Received | Injection delay (sec) |
|---|---|---|---|
| 06-0030 | placebo | naloxone | 230 |
| 07-0025 | naloxone | naloxone | 128 |
| 10-0001 | placebo | placebo | 149 |
| 11-0030 | naloxone | naloxone | 171 |
| 13-0004 | placebo | placebo | 374 |
| 14-0158 | placebo | placebo | 132 |

#### 4.3 Background and demographic characteristics

Baseline patient characteristics will be summarized using descriptive statistics (number, mean, standard deviation, median, minimum and maximum for quantitative measures and numbers and percentages for qualitative measures).
The following subjects' characteristics will be described in the population "inclusion":
- Centre
- Age and gender

The following subjects characteristics will be described in the population "mITT" to check the comparability of the naloxone group with the control group (descriptive statistics by group, no test):
Patients' characteristics

3537 - ENALEPSIE
- Centre
- Age and gender
- Body mass index
- Past epilepsy surgery
- Medical history
- Age at epilepsy onset (age – epilepsy duration)
- Epilepsy duration
- Epilepsy aetiology (including MRI results)
- Total number of seizures (i.e. focal seizures and GTCS) over the past three months
- Number of GTCS over the past three months
- Localization of the seizure onset zone, as assessed by video-EEG and/or video-SEEG data
- Number of anti- epileptic drugs (AED) at entry
- Comorbidity: depression, Sleep apnoea syndrome (SAS)
- Concomitant treatment: neurostimulation by vagus nerve stimulation (VNS), Serotonin reuptake inhibitors (SRI), Benzodiazepine
- Caffeine consumption
- AED withdrawal at randomisation
-Type of monitoring (scalp video-EEG or SEEG)
- Lateralization of the epileptogenic zone
- Localization of the epileptogenic zone (temporal vs extra-temporal)

Characteristics of the GTCS
- Seizure type (GCS type 1, 2 or 3)
- Duration of tonic phase
- Duration of tonic-clonic phase
- State of wakefulness (awake or sleep)
- Hypoxemia during the focal phase
- Early administration of 02
- O2 Administration (early or late)
- Post-ictal generalized EEG suppression (PGES)

#### 4.4 Analysis of the primary outcome

**Planned analysis before the unblinding**

This analysis will be performed on the population "mITT".
The primary endpoint calculation is based on an algorithm applied to spo2 records. In case of discrepancy with the investigator visual analysis, if the difference is less than ten seconds the algorithm result is preferred. Otherwise in case of artefact the manual result is preferred.
Decisions in blind review for ENALEPSIE study:
Patients 10-0008 and 14-0158 : value of algorithm

3537 - ENALEPSIE

Patients 10-0022 and 07-0025 : value of manual record

Patient 10-0064 : missing because no spo2

Decisions in blind review for REPOMSE study:

Patients 307, 434, 515, 557, 603, 788, 1052 and 1059 : value of algorithm

Patients 351, 475, 898, 926 and 1007 : value of manual record

Patients 102, 278 and 422 : missing because no spo2

##### 4.4.1 Formal analysis

**Modification to take into account the deviations to randomization after unblinding:**

- **The primary analysis**, i.e. Kaplan-Meier survival analysis to analyse the time to recovery of SpO2 ≥ 90% will not be performed in intention to treat, but using the actually received treatment. Because of the small sample size of the control group, no adjustment will be perform and patients in whom the study drug was administered > 120 seconds after the end of the seizure will not be excluded from the primary analysis. Only the 3 patients without GTCS and the 2 patients without SpO2 data will be excluded from the primary analysis. The analysis will compare the naloxone treatment to the placebo treatment using a bilateral log rank test at the 0.05 level of significance.

Also a time to event analysis (survival model) will be performed.

##### 4.4.2 Sensibility analysis

- **A secondary analysis of the primary outcome** will compare the naloxone treatment in ENALEPSIE patients to RESPOMSE patients used as an historic control group since the patients characteristics are similar.
  ENALEPSIE patients in whom the naloxone was administered > 120 seconds after the end of the seizure (3 patients in ENALEPSIE patients who received naloxone) will be excluded.
- **Potential factors for association with time to post-ictal recovery of O2 saturation** will be tested in a as explanation variables (listed in appendix). Naloxone treatment effect will be compare to the placebo treatment using a bilateral log rank test at the 0.05 level of significance.

In case of right censoring, a Kaplan meier estimation and a log rank test will be used. Also a time to event analysis (survival model) will be performed.

#### 4.5 Analysis of the secondary outcomes

**Planned analysis before the unblinding:**

**Modification to take into account the deviations to randomization after unblinding:**

The analyses will be restricted to two secondary endpoints:

- Total duration of the postictal coma, defined as the delay between the end of the seizure and the recovery of consciousness assessed by the ability to meet one single verbal command (handshake).
- Total duration of the postictal immobility, defined as the delay between the end of the seizure and the first spontaneous movement of the patient, as assessed on the video recording.

These analyses will compare the naloxone treatment in ENALEPSIE patients to the historic control group REPOMSE, using the same mehods than for the primary endpoint:

ENALEPSIE patients in whom the naloxone was administered > 120 seconds after the end of the seizure will be excluded.

Potential factors for association with time to postictal mobility and time to recovery of consciousness will be tested using the same variables than for the primary endpoint (listed in appendix).

#### 4.6 Data handling

##### 4.6.1 Management of missing data

Primary endpoint

Missing data for the patients for whom SPO2 is not recorded properly were discussed in blind review and cannot be replaced in the primary endpoint (mITT population).

Other endpoints

Missing data will not be replaced.

##### 4.6.2 Data transformation

No data transformation is planned

##### 4.6.3 Derived variables

The primary and secondary outcomes relative to pulse oximetry will be derived using an algorithm taking into account the values of SpO2 and the time of recorded values (hh:mm:ss). A special attention will be paid to artefacts as confusion factor.

#### 4.7 Subgroup analyses

No subgroup analyses are planned.

#### 4.8 Interim analyses

No interim analyses are planned.

#### 4.9 Statistical software

SAS ®version 9.4 or further (Copyright (c) 2002-2003 by SAS Institute Inc., Cary, NC, USA.).

R version 4.0.2 or further (R Core Team (2012). R: A language and environment for statistical computing. R Foundation for Statistical Computing, Vienna, Austria. ISBN 3-900051-07-0, URL http://www.R-project.org/.)

### 5 Quality

#### 5.1 Data

Controls will be provided in the data-management plan by the coordinating centre.

#### 5.2 Statistical analysis

Statistical analyses will be performed following the standard operating procedures of the Biostatistics department of the Hospices Civils de Lyon. The derivation of the primary endpoint will be compared to the investigator visual analysis. The results will be compared and errors will be discussed and corrected.

### 7 Appendix

#### 7.1 Potential factors for association with post-ictal recovery of O2 saturation

| Variable label | CRF table or external file | Association (Rheims et al 2019) | To include in SpO2 analysis | To include in population description | If not tested, reason to include in SpO2 analysis |
|---|---|---|---|---|---|
| Center | CRF (variable 1) | Not tested | | X | No placebo in some centers |
| Gender | CRF (variable 4) | No | X | X | |
| Age | Calculated (year of birth in file ENALEPSIE_Annees_de_naissance.xls) | No | X | X | |
| Epilepsy duration | Seizures & epilepsy | No | X | X | |
| Age at epilepsy onset | Seizures & epilepsy | No | X | X | |
| BMI | Past-history & comorbidities | No | X | X | |
| Number of anti-epileptic drugs (AED) at entry | Long term video scalp EEG monitoring | Not tested | | X | Only 2 patients treated with EAD |
| Treatment by VNS | Past & current treatment | Not tested | | X | Only 1 patient with VNS |
| PAST EPILEPSY SURGERY | Past & current treatment | Not tested | | X | No patient n the control group |
| Autres ATCD médicaux | Past-history & comorbidities | Not tested | | X | |
| Epilepsy aetiology (dont résultat IRM) | Seizures & epilepsy | Not tested | | X | |
| AED withdrawal at randomisation | CRF « Long term vidéo scalp… » or « Invasive EEG » | Not tested | | X | May be asked by reviewers |
| Treatment Benzodiazepine | Concomitant treatment | Not tested | Only 1 patient concerned in the control group | X | Suggested to be link to desaturation in Lacuey N, Martins R, Vilella L, et al. The association of serotonin reuptake inhibitors and benzodiazepines with ictal central apnea. Epilepsy Behav\textit{ }2019;98:73-79. |
| Type of video (scalp or SEEG) | « Long term video scalp » completed = scalp<br>« Invasive EEG » completed = SEEG | Not tested | | X | No potential association |

| | | | | | |
|---|---|---|---|---|---|
| Current depression | Past-history & comorbiditie | Not tested | X | **X** | May be a risk factor for desaturation (via dysfonction sérotoninergique) |
| Seizure type (generalised compulsive, focal, both) | VIDEO | No | **X** | **X** | Association with a confondant factor (Alexandre, Neurology 2015) |
| Duration of tonic phase | VIDEO | No | **X** | **X** | Usually associated in literature |
| Duration of tonic-clonic phase | VIDEO | No | **X** | **X** | - |
| State of wekefulness (awake or sleep) | VIDEO | No | **X** | **X** | Usually associated in literature |
| post-ictal generalized EEG suppression (PGES) | VIDEO | Yes | | X | Only for sclap, NA if SEEG |
| Localization of the Epileptogenic zone (temporal vs extra-temporal) | Long term video scalp EEG monitoring (lobe) | Yes | **X** | X | |

#### 7.2 Variables for endpoints

**Primary endpoint :**

- Delay to recovery of oxygen saturation calculated by the algorithm: Variable RETOUR
- Delay to recovery of oxygen saturation calculated with times given by the investigator visual analysis : Variable duree_hypox
- Primary endpoint calculated using one of the 2 variables: Variable primary

**Secondary Endpoints:**

o Total duration of the postictal generalized EEG suppression, This ouctome will only be assessed in patients undergoing scalp Video-EEG. Col AB Variable duree PGES (microvolts) -> 7 ou 8 else 0 if no PGES

o Total duration of the postictal coma, defined as the delay between the end of the seizure and the recovery of consciousness assessed by the ability to meet one single verbal command (handshake). Variable Duree_coma (table alldata)

o Total duration of the postictal immobility, defined as the delay between the end of the seizure and the first spontaneous movement of the patient: Variable Duree_immob table alldata
